# Case Study: Longitudinal immune profiling of a SARS-CoV-2 reinfection in a solid organ transplant recipient

**DOI:** 10.1101/2021.03.24.21253992

**Authors:** Jonathan Klein, Anderson F. Brito, Paul Trubin, Peiwen Lu, Patrick Wong, Tara Alpert, Mario A. Peña-Hernández, Winston Haynes, Kathy Kamath, Feimei Liu, Chantal B. F. Vogels, Joseph R. Fauver, Carolina Lucas, Jieun Oh, Tianyang Mao, Julio Silva, Anne L. Wyllie, M. Catherine Muenker, Arnau Casanovas-Massana, Adam J. Moore, Mary E. Petrone, Chaney C. Kalinich, Yale IMPACT Research Team, Charles Dela Cruz, Shelli Farhadian, Aaron Ring, John Shon, Albert I. Ko, Nathan D. Grubaugh, Benjamin Israelow, Akiko Iwasaki, Marwan M. Azar

## Abstract

Prior to the emergence of antigenically distinct SARS-CoV-2 variants, reinfections were reported infrequently - presumably due to the generation of durable and protective immune responses. However, case reports also suggested that rare, repeated infections may occur as soon as 48 days following initial disease onset. The underlying immunologic deficiencies enabling SARS-CoV-2 reinfections are currently unknown. Here we describe a renal transplant recipient who developed recurrent, symptomatic SARS-CoV-2 infection - confirmed by whole virus genome sequencing - 7 months after primary infection. To elucidate the immunological mechanisms responsible for SARS-CoV-2 reinfection, we performed longitudinal profiling of cellular and humoral responses during both primary and recurrent SARS-CoV-2 infection. We found that the patient responded to the primary infection with transient, poor-quality adaptive immune responses. The patient’s immune system was further compromised by intervening treatment for acute rejection of the renal allograft prior to reinfection. Importantly, we also identified the development of neutralizing antibodies and the formation of humoral memory responses prior to SARS-CoV-2 reinfection. However, these neutralizing antibodies failed to confer protection against reinfection, suggesting that additional factors are required for efficient prevention of SARS-CoV-2 reinfection. Further, we found no evidence supporting viral evasion of primary adaptive immune responses, suggesting that susceptibility to reinfection may be determined by host factors rather than pathogen adaptation in this patient. In summary, our study suggests that a low neutralizing antibody presence alone is not sufficient to confer resistance against reinfection. Thus, patients with solid organ transplantation, or patients who are otherwise immunosuppressed, who recover from infection with SARS-CoV-2 may not develop sufficient protective immunity and are at risk of reinfection.

## Introduction

The dynamics and duration of adaptive immune responses to SARS-CoV-2 infection have been described in association with disease severity and the rate of viral clearance, yet the correlates of adaptive immunity responsible for preventing reinfection remain incompletely characterized. In studies of SARS-CoV-2 infection in animal models (mice^1,2^, hamsters^3,4^, and rhesus macaques^5–8^), both vaccine-induced and natural infection-induced immunity are sufficient for protection from SARS-CoV-2 rechallenge. Recent Phase III vaccine clinical trials^9^, as well as epidemiologic studies of natural infection^10^, have also demonstrated robust development of protective immunity in humans. Taken together, these data unambiguously demonstrate that adaptive immunity confers protection against SARS-CoV-2 infection in the majority of cases.

However, rare case reports of SARS-CoV-2 reinfection by antigenically similar variants have also been documented as soon as 48 days from primary symptom onset^11–18^ (**Extended Data Table 1**). Whether these reinfections are the direct result of deficient adaptive immune responses to the primary infection, or are the result of waning adaptive immunity, is currently unknown. Notably, cases of persistent SARS-CoV-2 infection among patients with underlying genetic defects (such as X-linked agammaglobulinemia^19^) or acquired defects (B cell depletion therapy^20^) in humoral immunity have been reported. Immunocompromised COVID-19 patients with prolonged infection also achieved viral clearance when treated with multiple doses of convalescent plasma^21^, demonstrating the sufficiency of humoral responses in clearing SARS-CoV-2 infection. The role of cellular immunity in protection from SARS-CoV-2 infections is also a subject of intense investigation. Studies demonstrate that most COVID-19 patients develop SARS-CoV-2 specific CD4^+^ and CD8^+^ T cells^22^ and reports of durable T-cell memory responses to related SARS-CoV-1 infection lasting up to 17 years after initial infection^23^. While neutralizing antibodies are a correlate of protection, non-human primate models have demonstrated reduced virological control in the upper respiratory tract in CD8^+^ depleted convalescent animals upon reinfection^24^, suggesting that both arms of the adaptive immune response may be required for optimal clearance and protection against SARS-CoV-2 infection.

Due to the rarity and complexity involved in investigation of human SARS-CoV-2 reinfections, complete immune profiles exploring the magnitude and extent of these adaptive immune responses in paired primary infection and reinfection are lacking. Identifying the deficient features of initial adaptive immune responses that enables subsequent SARS-CoV-2 reinfection will help to further define the correlates of immune protection in humans.

## Results

### Clinical presentation of immunocompromised solid organ transplant recipient with SARS-CoV-2 reinfection

In March 2020, a man between 60-70 years old residing in a transitional group living facility with a medical history notable for end-stage renal disease, for which he had undergone living-donor renal transplantation two years prior, was hospitalized with fevers, fatigue, and dry cough (**Fig. 1**). Induction immunosuppression for renal transplantation had consisted of antithymocyte globulin, while maintenance immunosuppression initially included tacrolimus (a calcineurin inhibitor that inhibits T-cell cytokine production), mycophenolate mofetil (MMF, a B and T lymphocyte anti-proliferative agent), and low-dose prednisone. By the time of hospitalization, belatacept (a T lymphocyte costimulation blocker) had been substituted for tacrolimus due to the development of calcineurin-induced neurotoxicity, and prednisone had been discontinued due to perceived exacerbation of psychiatric illness. Persistent neutropenia complicated the post-transplantation course, requiring substitution of prophylactic inhaled pentamidine for trimethoprim-sulfamethoxazole and frequent infusions of filgrastim. Upon hospitalization, SARS-CoV-2 infection was diagnosed via reverse-transcriptase polymerase chain reaction (RT-PCR) performed on a nasopharyngeal swab (NP) specimen. He was subsequently enrolled in the Yale Implementing Medical and Public Health Action Against Coronavirus CT (IMPACT) study, a biospecimen repository housing clinical and demographic data as well as respiratory, blood, and other tissue samples from patients with confirmed COVID-19 at Yale New Haven Hospital. He developed symptomatic moderate COVID-19 for which he received hydroxychloroquine and atazanavir for 5 days and a single dose of tocilizumab at 8 milligrams/kilogram (mg/kg). MMF was paused and a reduced dose of belatacept was administered in the setting of acute infection. The oxygen requirement peaked at 4 liters per minute by nasal cannula; by 13 days from symptom onset (DFSO), the patient was transitioned to room air. Though the patient was asymptomatic thereafter, nasopharyngeal (NP) swabs and saliva (SL) from the patient remained positive for SARS-CoV-2 by PCR throughout the hospital stay (**Extended Data Table S2**). The patient was discharged from the hospital on 27 DFSO to the transitional group residential facility after a 14-day period without hypoxia, reemergence of symptoms, or other clinical signs of infection. MMF was restarted on discharge.

**Figure 1.**
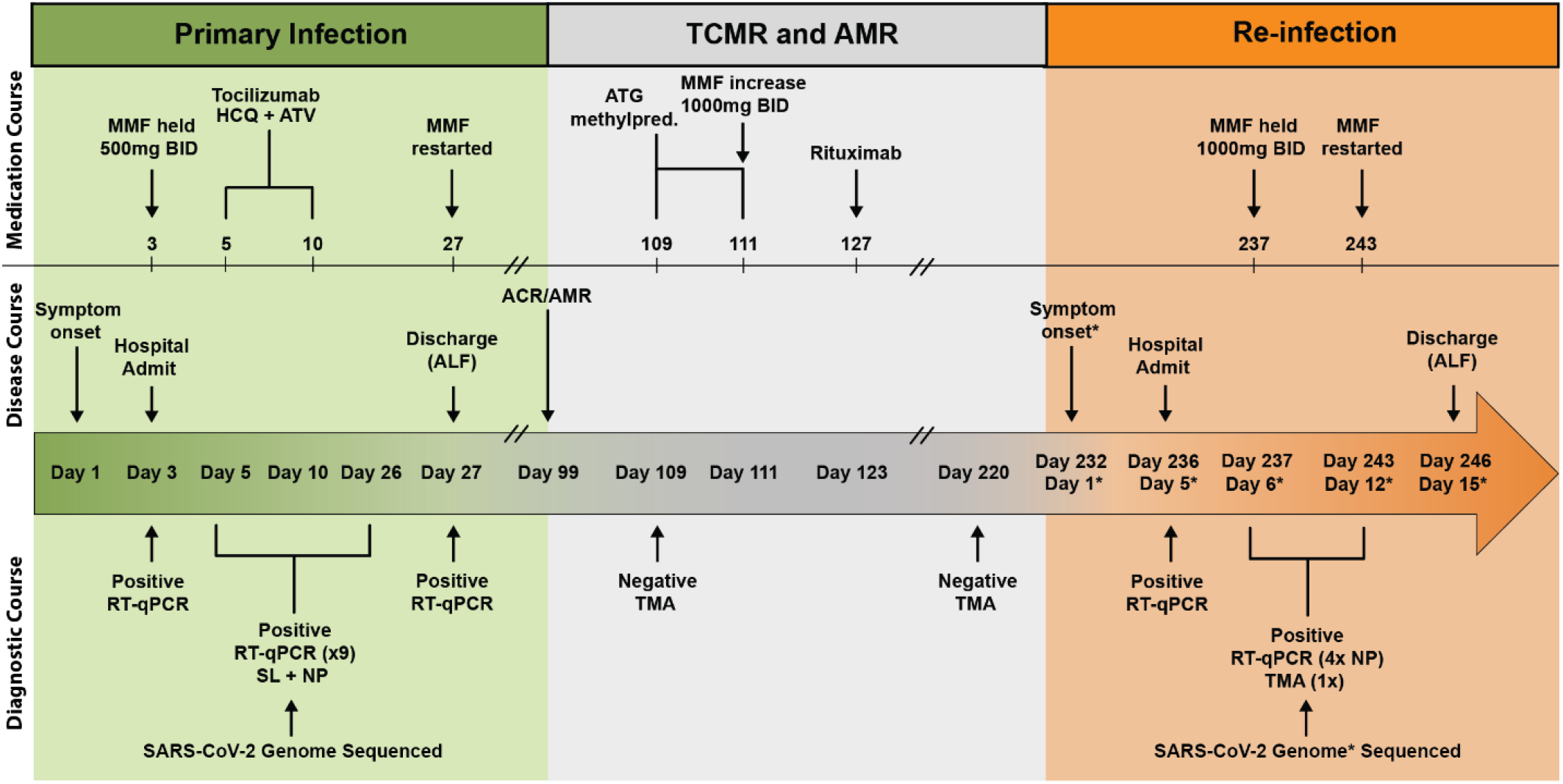
SARS-CoV-2 reinfection clinical timeline. Summary of patient’s disease course divided into distinct clinical periods: primary infection (green), graft rejection and immunosuppressive therapies (grey), and SARS-CoV-2 reinfection (orange). Clinical annotations are stratified into rows based on content (Medications, Disease Course, Diagnostic Testing). Arrows indicate specific events, brackets indicate duration of treatment or testing where applicable. Asterisks (*) indicate annotations specific to SARS-CoV-2 reinfection. Double line breaks (/ /) indicate condensing of clinical timeline for display. Abbreviations (Top to bottom, left to right): T cell mediated rejection (TCMR); antibody-mediated rejection (AMR); mycophenolate mofetil (MMF); BID (twice daily); hydroxychloroquine (HCQ); atazanavir (ATV); anti-thymocyte globulin (ATG); methylprednisolone (methylpred.); assisted living facility (ALF); Real-time quantitative polymerase chain reaction (RT-qPCR); saliva (SL); nasopharyngeal (NP); transcription mediated amplification (TMA).

Approximately 10 weeks after discharge, a kidney allograft biopsy was performed because of increasing serum creatinine and was notable for evidence of acute T-cell-mediated rejection (TMCR) and antibody-mediated rejection (AMR) of the transplanted organ (**Fig. 1**). He was readmitted and treated with 400 mg of antithymocyte globulin and 125 mg of methylprednisolone. Belatacept was continued, low-dose prednisone was restarted, and the MMF dose was increased. Notably, a nasopharyngeal swab collected at the time was negative for SARS-CoV-2 using a non-quantitative transcription-mediated amplification (TMA) test. The patient remained asymptomatic and was discharged back to the transitional living facility. He received rituximab 1 week after discharge to address AMR.

Approximately 15 weeks after this hospitalization, the patient underwent repeat renal allograft biopsy for evaluation of polyomavirus-associated nephropathy that demonstrated evidence of mild remnant AMR (**Fig. 1**). Ongoing neutropenia necessitated additional infusions of filgrastim. At 220 DFSO, a NP swab collected from the patient was again negative for SARS-CoV-2 using TMA.

Approximately 4 months after the diagnosis of rejection and 7 months from his primary COVID-19 diagnosis, the patient was readmitted to the hospital with fatigue and nonproductive cough (**Fig. 1**). Repeat SARS-CoV-2 PCR of NP samples returned positive at 236 DFSO / 5 days from reinfection symptom onset (DFSO*) with cycle thresholds to targets N1 and N2 of 27.34 and 27.15, respectively. The patient did not develop fevers or hypoxia, had no evidence of pneumonia on chest imaging, and did not require COVID-19-specific therapy. SARS-CoV-2 IgG was reactive at 5 DFSO*. Isolation precautions were reinstituted for the 10-day duration of hospitalization and were maintained after his return to the group living facility.

### Genome sequencing reveals two distinct lineages of SARS-CoV-2 during primary infection and reinfection

Following symptom onset during the primary infection in March 2020, both nasopharyngeal and saliva specimens tested positive by PCR, and nasopharyngeal specimens were whole genome sequenced for phylogenetic analysis^25^. Additional nasopharyngeal and saliva specimens were collected and sequenced during the reinfection episode in November 2020 (**Extended Data Table 2**). To rule out the possibility of persistent SARS-CoV-2 infection, which has been previously reported^26–29^, we compared the virus genomes sequenced from specimens collected 7 DFSO in the primary infection (NP swab), and 5 DFSO* during the reinfection (NP swab and saliva). Phylogenetic analysis revealed that viruses from the primary infection and reinfection belong to 2 distinct clades within the SARS-CoV-2 lineage B: clade B.1 in the primary infection in March 2020, and B.1.280 in the reinfection in November 2020 (**Fig. 2a**). Specifically, the virus genome sequenced from the reinfection (**Fig. 2a, c (green)**) had 12 mutations not observed in the virus sequenced from the primary infection (**Fig. 2c (orange)**): 4 synonymous and 8 non-synonymous. Among the mutations that alter amino acid identity relative to the SARS-CoV-2 reference genome (Wuhan-Hu-1, GenBank: MN908947), both viruses expressed the spike protein with glycine in position 614 (D614G), but only the virus from the reinfection had an additional polymorphism at spike A1078S, close to the transmembrane connector domain in the S2 subunit^30^ (**Fig. 2b; Extended Data Fig. S2**). Importantly this mutation is not located within the SARS-CoV-2 spike receptor binding domain, which is the primary target of neutralizing antibodies (**Extended Data Fig. S2**), nor has it been reported among SARS-CoV-2 variants of concern (VOC) B.1.1.7, B.1.351, or P.1 that display variable evasion of humoral immune responses^31^.

**Figure 2.**
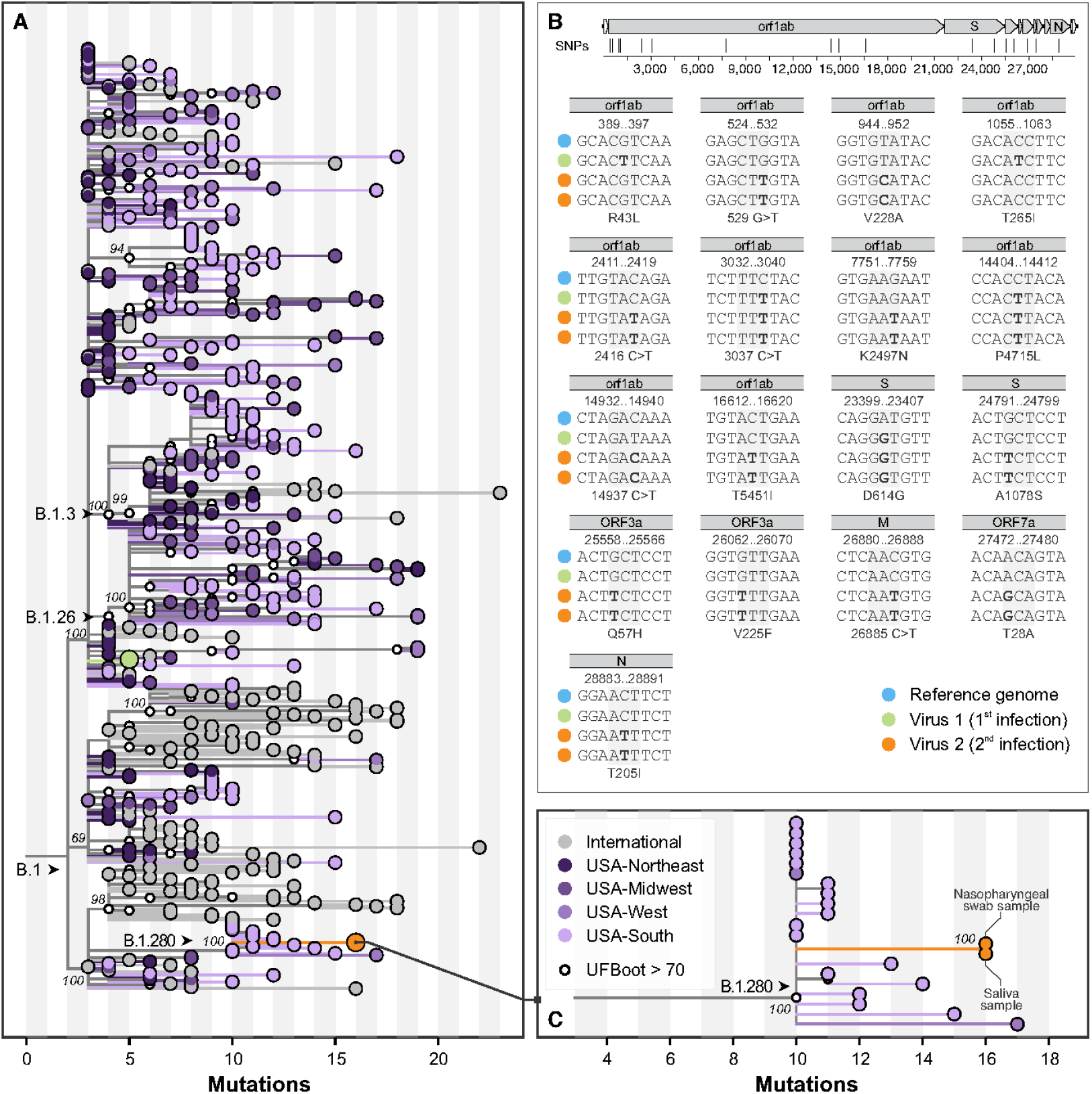
Maximum likelihood phylogeny of SARS-CoV-2 whole genomes. **(A)** Global tree showing the evolutionary relationship of 561 lineage B.1 SARS-CoV-2 genomes, including three samples from the patient’s two independent infections as described in this study (green, primary infection; orange, reinfection). These viruses belong to two sublineages, which evolved independently of each other since their most recent common ancestors, which circulated in the Northeast United States in March 2020. **(B)** Profile of mutations observed in genomes from both infections, compared with the reference genome (GenBank: MN908947), shown at the top, highlighting the positions of the SNPs shown in the panel. Highlights of the two genomic sequences obtained from the reinfection are shown. **(C)** A zoomed view of the clade demonstrating relatedness of viruses in the reinfection group reveals their relatedness to viruses that circulated in the Southern United States (state of Florida), the likely origin of that sublineage.

Our phylogenetic analysis also demonstrates that the distinct viral lineages identified from the patient’s primary infection and subsequent reinfection diverged from their common ancestor around March 2020 (**Fig. S1**), suggesting intra-host evolution in the setting of persistent infection to be an unlikely explanation for this case and providing unambiguous evidence of reinfection. To rule out the remote possibility of the presence of multiple SARS-CoV-2 lineages during reinfection, we also sequenced virus genomes from both saliva and nasopharyngeal swabs collected during the reinfection (**Fig. 1c**) and found them to be identical. Lastly, we analyzed the geographic distributions of circulating SARS-CoV-2 lineages and discovered that the sub-lineage of viruses identified in the reinfection likely first circulated in the Southern US in June of 2020 before being reintroduced to the Northeast US. This patient’s primary residence is located within the Northeast, and he reported no travel since discharge from the primary SARS-CoV-2 infection in March of 2020 (**Fig. S1**), confirming that his SARS-CoV-2 reinfection was likely the result of a broad geographic reintroduction and unlikely to represent an instance of persistent SARS-CoV-2 infection.

Given our findings of the distinct genetic lineages of each SARS-CoV-2 isolate, the lack of multiple strains of SARS-CoV-2 during reinfection, and the congruent geographic patterns of the patient’s clinical narrative, we established that our case represents a genetically confirmed SARS-CoV-2 reinfection and next sought to identify the specific immune correlates that conferred this susceptibility.

### Immunologic profiling reveals naive lymphocyte depletion and poor humoral immunity

During the patient’s primary SARS-CoV-2 infection, we performed longitudinal whole blood sampling which was separated into peripheral blood mononuclear cells (PBMC) and serum fractions at 7, 15, and 23 DFSO. PBMCs were analyzed by multidimensional flow cytometry and serum was analyzed with multiplex ELISA to measure 71 cytokines (**Fig. 3-4; Extended Data Fig. S3-S4**).

**Figure 3.**
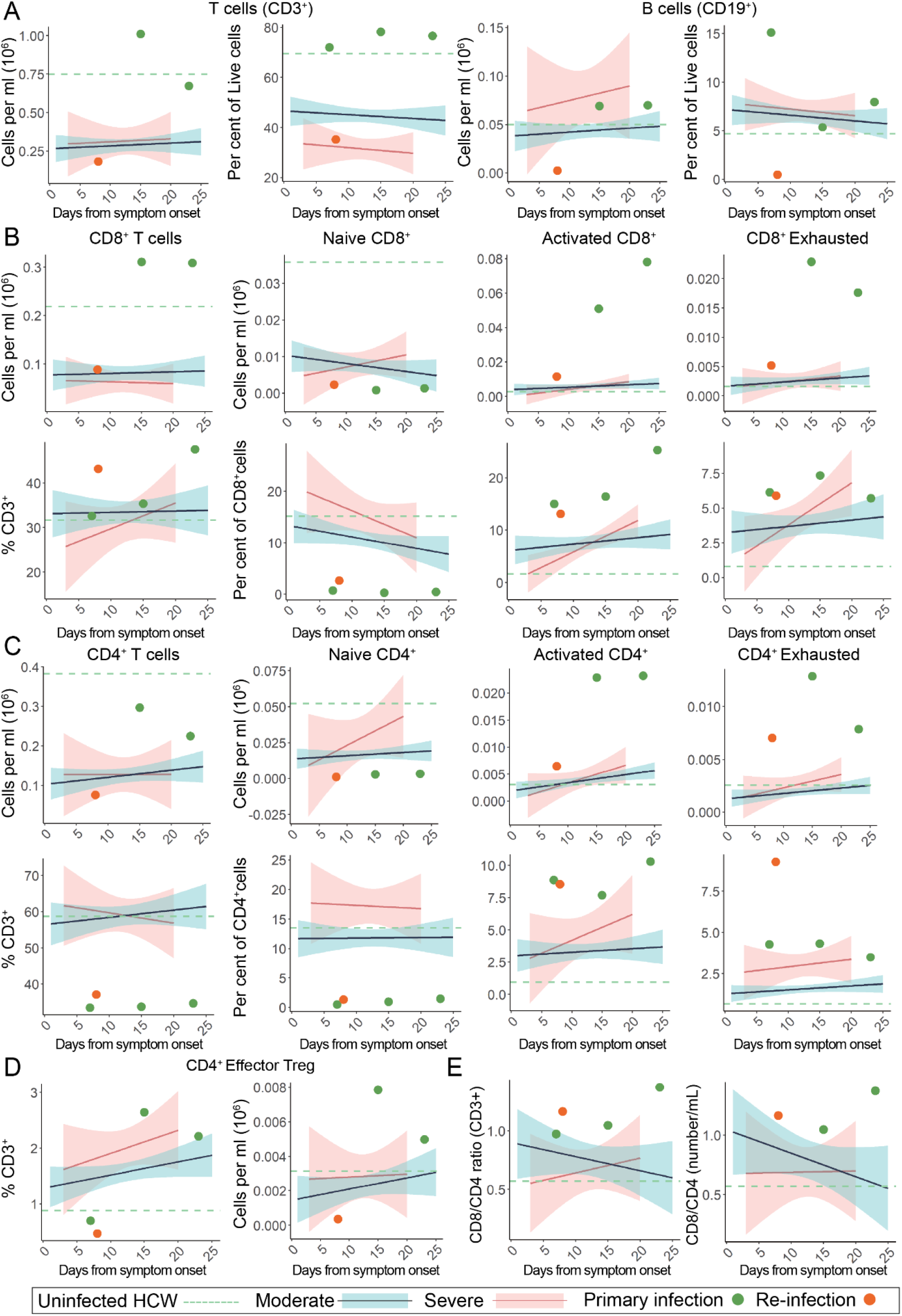
Peripheral lymphocyte profiling of SARS-CoV-2 primary and reinfection demonstrates persistent T cell exhaustion and loss of B cells. For all graphs, blue linear least squares regression lines and corresponding shading represent the average trend and error bars, respectively, for patients with moderate COVID-19. Red linear least squares regression lines and corresponding shading represent the average trend and error bars, respectively, for patients with severe COVID-19. The dashed green line represents the average value of healthy, uninfected healthcare workers (HCW) plotted as a constant value across all days for reference. Individual scatter points represent the values for the patient during the primary SARS-CoV-2 infection (green) at 7, 15, and 23 DFSO and the reinfection (orange) at 8 DFSO*. **(A)** Total T cells and B cells isolated from patient whole blood. **(B)** CD8^+^ T cell subsets plotted as number (top) and relative percentage of parent (bottom). **(C)** CD4^+^ T cell subsets plotted as number (top) and relative percentage of parent (bottom). **(D)** CD4^+^ T_FH_ cell subsets plotted as number and percentage of parent CD3^+^. **(E)** CD8^+^ / CD4^+^ ratios calculated relative to days from symptom onset.

In comparison to disease severity and DFSO matched patients from our larger IMPACT cohort, we found that the patient differed significantly in both immune cell subtype composition as well as cytokine expression during his primary infection. Notably, during the primary SARS-CoV-2 infection, the patient maintained very high levels of circulating T-cells and did not suffer from a T-cell lymphopenia as is characteristic of symptomatic COVID-19 patients^32^ (**Fig. 3a**). Not only was general lymphopenia absent, there also was no specific loss of CD8^+^ T cells, as can be seen in more severe cases of COVID-19^33,34^ (**Fig. 3b**). Importantly the patient also demonstrated a relatively higher, rather than characteristically depressed, CD8^+^/CD4^+^ ratio primarily as a result of his diminished CD4^+^ populations. (**Fig 3b-c**,**e; Extended Data Fig S3b**). With regards to functionality, the patient’s CD8^+^ and CD4^+^ T-cells exhibited broad increases activation markers (CD38^+^, HLA-DR^+^), exhaustion/terminal differentiation markers (PD1^+^, TIM-3^+^), and effector T regulatory cell markers (PD1^+^, TIM-3^+^, CD25^+^, CD127^-^, HLA-DR^+^) (**Fig. 3b-d**). In comparison to the larger IMPACT cohort, this patient’s immunological profile was uncharacteristic of either moderate or severe SARS-CoV-2 infection, and instead resembled an immunophenotype consistent with chronic antigen exposure. Importantly, we found that the patient also had very low numbers of circulating naïve CD4^+^ and CD8^+^ T cells at the time of primary SARS-CoV-2 infection, which are required for the generation of potent de novo antiviral response.

To assess whether alterations in immune cell composition contributed to reinfection, we again performed multidimensional flow cytometry on PBMCs isolated from the patient at 5 DFSO* (**Fig. 3a-c**, orange). In comparison to results from the primary infection (**Fig. 3a-c**, green), we found a general loss of circulating lymphocytes, while myeloid cell subsets remained at similar levels as seen during his primary infection (**Extended Data Fig. 3S**). We suspect that this broad depletion of lymphocytes was due to intervening treatment with antithymocyte globulin and rituximab during an episode of graft rejection 3 months prior to his reinfection (**Fig. 1**). It is also possible that the SARS-CoV-2 reinfection exacerbated this global depletion, although decreases in circulating B cell populations have not been widely reported in COVID-19 patients. Among the patient’s remaining T cell populations, and in the context of recent anti-rejection treatment, the patient again presented with largely depleted pools of naïve CD4^+^ and CD8^+^ T-cells and with continued activation and exhaustion among effector CD4^+^ and CD8^+^ T cell populations. In contrast to the primary infection, we found an almost complete depletion of CD19^+^ B cells, likely as a result of intervening rituximab treatment (**Fig. 3a**). Consistent with our findings during the primary infection, the patient again presented with an immunophenotype suggestive of chronic antigen engagement, but with globally reduced lymphocytes likely by the treatment for TMCR and AMR.

To investigate the full extent of immunological dysfunction present in the patient during the primary SARS-CoV-2 infection, we next explored whether altered cytokine signaling could have contributed to the patient’s poor initial adaptive immune response. Accordingly, we performed multiplex cytokine analysis from the patient’s serum and found that the patient had globally elevated cytokines (**Extended Data Fig. S4**) including IL-10, IFNα, IFNλ, IL-1α, TNFα, TRAIL, and IL-27 at all sampled points during primary infection. Other markers of T-cell functionality, including secreted cytokine IFNγ and T cell activating cytokines IL-18 and IL-12, remained elevated through the patient’s course of infection even after improvement in COVID-19 symptoms (**Fig. 4a**). In contrast to this patient, a disease-severity matched COVID-19 cohort showed either no elevation, or conversely, a reduction in levels of these cytokines over their course of infection. Additionally, the patient’s IL-15 and IL-7 levels, required for maintenance of naïve T-cell pools, were also persistently elevated (**Fig. 4b**). These data suggest that persistent utilization of T-cell populations - likely a result of continual immunological response to the patient’s allograft - rather than poor production of cytokines may be responsible for low numbers of naïve T cells at the time of primary SARS-CoV-2 infection.

**Figure 4.**
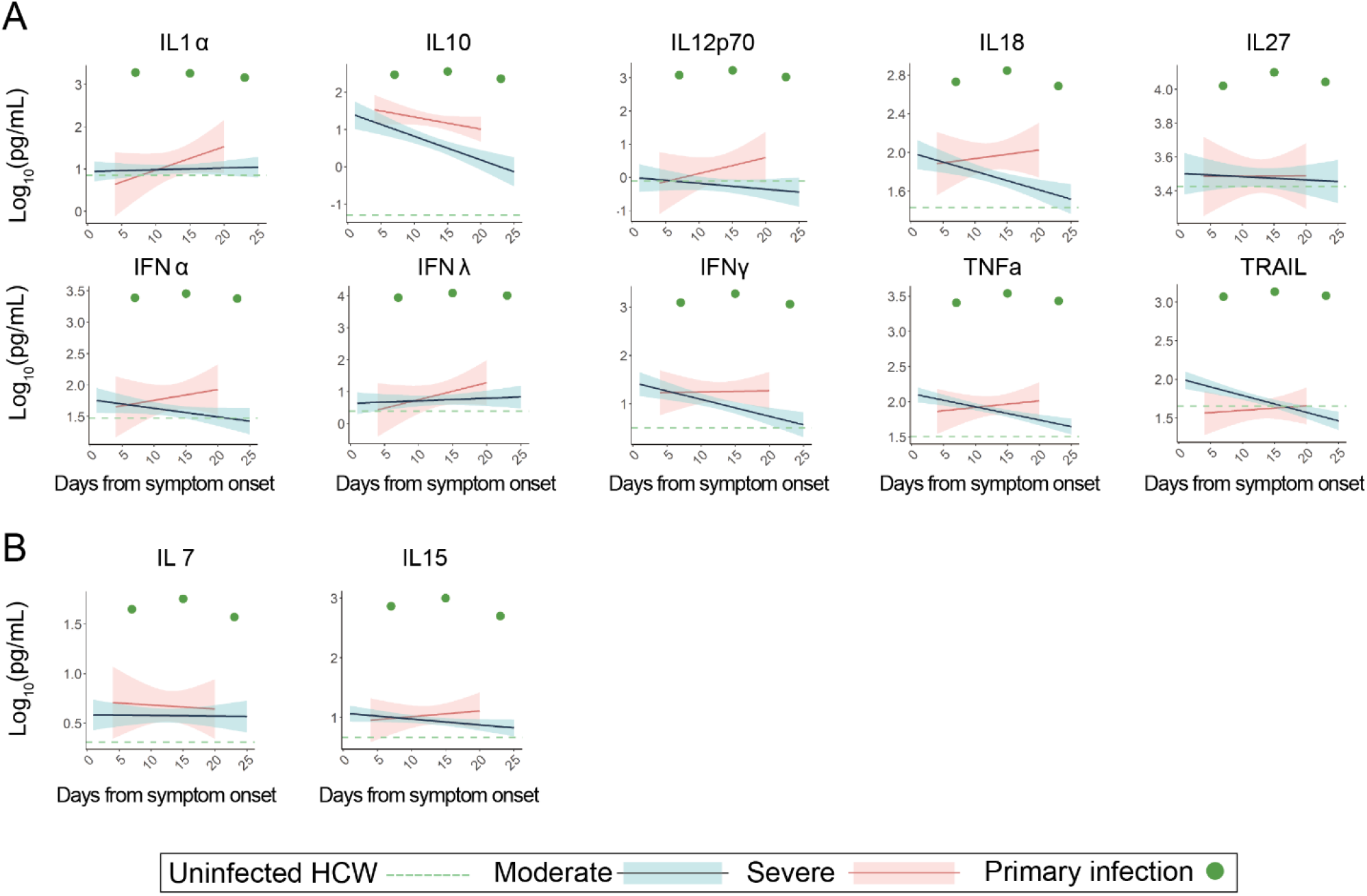
Peripheral cytokine profiling demonstrates broad increases in activation markers suggestive of chronic immune engagement. For all graphs, blue linear least squares regression lines and corresponding shading represent the average trend and error bars, respectively, for patients with moderate COVID-19. Red linear least squares regression lines and corresponding shading represent the average trend and error bars, respectively, for patients with severe COVID-19. The dashed green line represents the average value of healthy, uninfected healthcare workers (HCW) plotted as a constant value across all days for reference. Individual scatter points represent the values for the patient during the primary SARS-CoV-2 infection (green) at 7, 15, and 23 DFSO. **(A)** Serial measurements of various cytokines plotted against days from symptom onset **(B)** Select cytokines responsible for naive T-cell proliferation and maintenance

Given the patient’s loss of B cells prior to reinfection following the administration of rituximab (**Fig. 1, Fig. 3**), we initially hypothesized that SARS-CoV-2 reinfection may have also been the result of loss of humoral immunity. Accordingly, we first assessed anti-SARS-CoV-2 IgG and IgM levels by ELISA during primary infection and found that the patient produced typical levels of SARS-CoV-2 specific antibodies (S1 and RBD) compared with other hospitalized COVID-19 patients (**Fig. 5a**). Increasing S1 IgG and IgM levels positively correlated with rising RT-qPCR CT values specific for SARS-CoV-2 genomes (*i*.*e*. decreasing viral load), suggesting their role in resolution of the primary SARS-CoV-2 infection. During the patient’s reinfection, and in the setting of few circulating B-cells, we found an accelerated S1 IgG response that was again positively correlated with RT-qPCR CT values, suggestive of a memory response upon pathogen rechallenge (**Fig. 5b-c**). Moreover, there was a complete absence of S1 specific IgM during reinfection, consistent with a memory response to SARS-CoV-2 infection (**Fig. 5b**).These results suggest that antiviral antibodies were not lost during rituximab treatment as initially hypothesized, and furthermore that the source of S1-specific IgG during the reinfection was likely due to long-lived plasma cells (which are not depleted by rituximab^35^) generated during initial SARS-CoV-2 infection rather than a de novo response to the reinfection.

**Figure 5.**
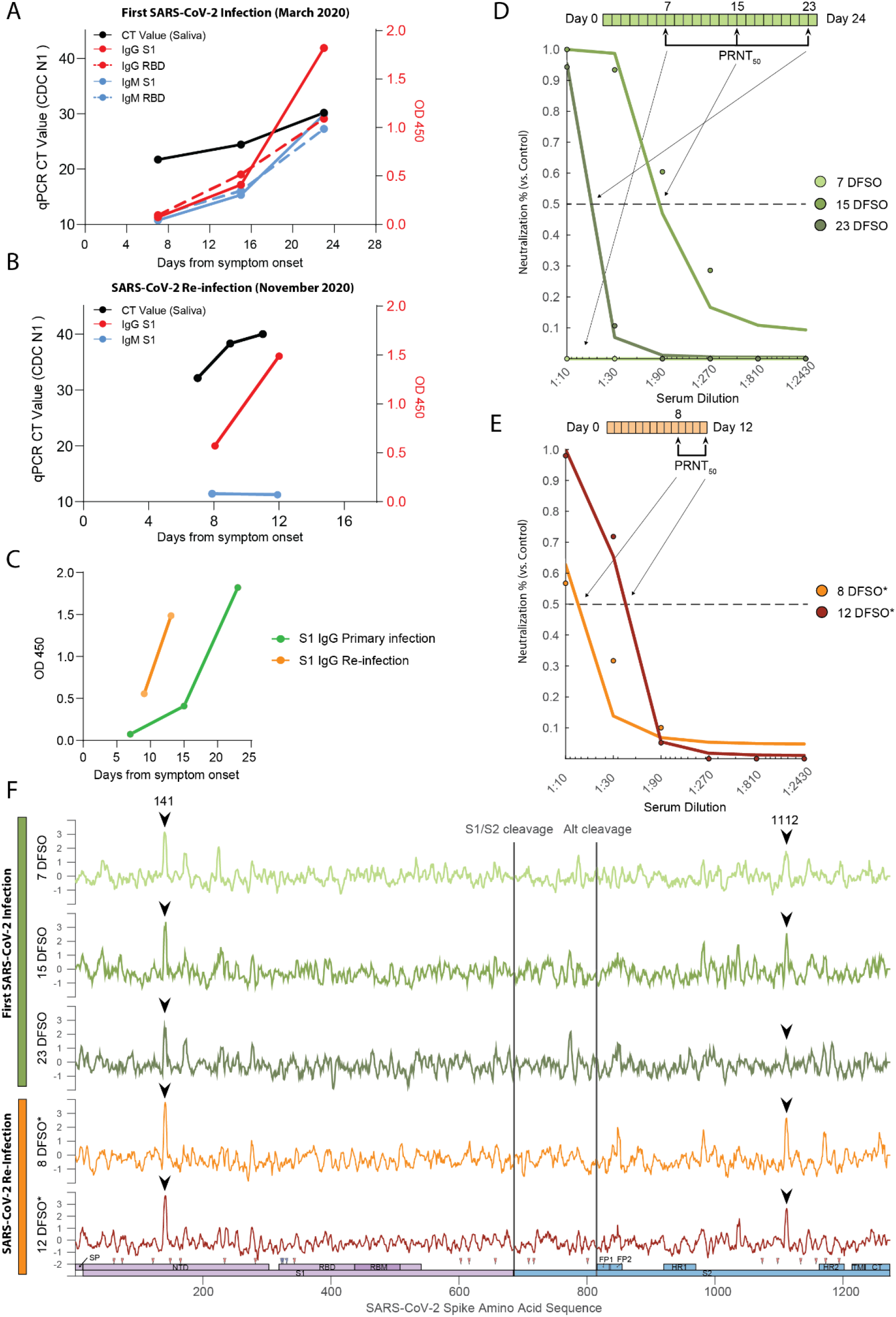
Humoral responses to primary and recurrent SARS-CoV-2 infection. **(A)** SARS-CoV-2 spike S1 (S1) and receptor binding domain (RBD) ELISA values, measured at optical density 450 (O.D. 450), as observed during the patient’s primary SARS-CoV-2 infection. Values are plotted against days from symptom onset (DFSO). Cycle threshold (CT) RT-qPCR values are shown in black (left y-axis) and corresponding ELISA data is shown in red (IgG) and blue (IgM) (right y-axis, red). Solid lines correspond to S1, dashed lines correspond to RBD **(B)** S1 ELISA values from the patient’s SARS-CoV-2 reinfection in November of 2020. CT qPCR values are shown in black (left axis) and corresponding ELISA data is shown in red (IgG) and blue (IgM) (right axis). Values are plotted against reinfection days from symptom onset (DFSO*) **(C)** ELISA S1 IgG trajectories plotted from primary and reinfection **(D, E)** Longitudinal PRNT_50_ assays for each sample collected during the patient’s SARS-CoV-2 primary (top, green) and reinfection (bottom, orange) episodes. Dots represent neutralization of the patient’s serum relative to healthy, uninfected healthcare workers. Solid lines represent the best fit of a generalized linear model for estimating serum IC_50_ values. Condensed clinical timeline (above) shows timing of PRNT50 assays relative to days from symptom onset. **(F)** PIWAS tiling data representing binding locations of patient’s antibodies against Spike protein. Samples are ordered longitudinally by rows (primary infection (green); reinfection (orange)) to track humoral dynamics. Shared peaks and respective peak heights between the primary and reinfection are annotated (black arrows). A map of SARS-CoV-2 spike domains is provided for reference against antibody binding locations (bottom).

To assess the neutralizing capacity of anti-SARS-CoV-2 antibodies present during both the primary and recurrent SARS-CoV-2 infections, we performed longitudinal PRNT50 assays and calculated the corresponding serum dilution IC50 values for each time point (**Fig. 5d-e**). While the patient developed neutralizing antibodies by 15 DFSO, they were transient in nature and significantly declined in potency by 23 DFSO. This atypical neutralizing antibody response is not consistent with other large-scale studies that show persistence of neutralization capacity following SARS-CoV-2 infection (t1/2=90 days; 95% CI: 70-125 days). Furthermore, the neutralization capacity was notably reduced even in comparison to other hospitalized COVID-19 patients of matched disease severity (**Extended Data Fig. S5a-b)**. Longitudinal analysis of serum samples was not performed during the intervening period between primary infection and reinfection; however, early hospital clinical laboratory serologic assays showed persistence of anti-SARS-CoV-2 IgG at 5 DFSO*. We were able to assess neutralizing antibodies during the reinfection, and found that neutralizing antibodies were present at 8 DFSO* and increased slightly by 12 DFSO*. Similar to the primary infection they were of poor neutralizing capacity relative to other COVID-19 patients (**Fig S5a; Extended Data Fig. S6**). Given that the patient was depleted of naive circulating B cells, had no IgM response, and had detectable circulating antibodies as early as 5 DFSO*, we hypothesized that these neutralizing antibodies observed during the reinfection reflected antibodies generated from the primary infection, rather than a new humoral response to the reinfection. To examine whether these neutralizing antibodies targeted the same regions with in the SARS-CoV-2 spike protein, we performed linear epitope mapping of this patient’s antibody binding using Serum Epitope Repertoire Analysis (SERA) - a random bacterial display peptide library - coupled with a recently described bioinformatic method that enriches for antigen-specific antibody binding signals relative to healthy (uninfected) controls (Protein-based Immunome Wide Association Study, “PIWAS”)^36^. Using this approach, we found two characteristic PIWAS peaks - signifying locations of peak patient antibody binding - at identical locations in both the primary infection and reinfection (**Fig. 5f**, black arrows). These peaks of antibody binding were centered on amino acid 141 in the N-terminal domain of S1 and on amino acid 1112 in the S2 domain of Spike. The high degree of concordance in peak locations between primary infection and reinfection suggests the same antibody-secreting population responded to both infections. Importantly, this peak is distinct from the Spike amino acid mutation at 1078 that was found only in the reinfection isolate (**Extended Data Fig. S2**), suggesting that viral evasion of the antibody response generated during the primary infection was unlikely to be responsible for reinfection.

In summary, we found that the patient developed an antigen-specific, neutralizing antibody response during his primary SARS-CoV-2 infection; that this neutralizing antibody response likely developed into a long-lived plasma cell population; and that it was insufficient to provide protection against reinfections with a novel lineage of SARS-CoV-2 that bore no evidence of viral immune evasion.

## Discussion

We have described a case of symptomatic SARS-CoV-2 reinfection in a solid organ transplant recipient and profiled the unique immunological dysfunctions present during both initial SARS-CoV-2 infection and reinfection. Through extensive clinical investigation and phylogenetic analysis of virus sequences, we confirmed that the patient was reinfected with a genetically distinct lineage of SARS-CoV-2, which was neither the result of persistent infection nor the result of infection by an antigenically distinct SARS-CoV-2 variant. Accordingly, we investigated the potential mechanistic causes of this patient’s multiple SARS-CoV-2 infections by performing longitudinal immunologic profiling during both initial SARS-CoV-2 infection and reinfection.

Multiple recent longitudinal studies have shown that the majority of COVID-19 patients, even those with mild or asymptomatic infection, develop long-lasting SARS-CoV-2 specific cellular and humoral adaptive immunity for as long as 8 months^37^. In contrast, a series of case reports and this manuscript report that SARS-CoV-2 reinfections can occur between 48 and 236 days from initial infection (**Extended Data Table 1**). The discrepancy between frequent, durable protective immune responses generated during most SARS-CoV-2 infections and rare cases of reported reinfection by antigenically similar variants is currently unexplained. While the protective capacity of humoral responses against SARS-CoV-2 infection is apparent, large variability in magnitude of responses between patients has been shown in multiple longitudinal studies. The underlying immunologic correlates of this variability, and ultimately what is required to develop strong humoral responses, have not been fully elucidated.

In this case study, we found that a failure of humoral immunity may have led to this patient’s SARS-CoV-2 reinfection. We investigated both the dynamics of antibody production as well as the general quality of antibodies produced by the patient. Our initial analysis of humoral responses indicated that the patient mounted a typical IgM and IgG response to SARS-CoV-2 primary infection, as assessed by longitudinal ELISA measurements. While total anti-SARS-CoV-2 antibody production was not particularly hampered in this patient, the neutralizing antibody response was clearly defective.

To address the underlying cellular defects that may have led to a poor neutralizing antibody response, we performed flow cytometry analysis of PBMC populations during primary infection and reinfection. We found significant differences in the patient’s T cell composition and phenotype relative to other patients with COVID-19. While T cell lymphopenia is common in even mild cases of COVID-19 and a characteristic response to many other viral infections, this patient did not develop T cell lymphopenia, and but instead presented with a profound and relatively specific reduction in naive T cell pools, most significantly in their CD4 compartment. Reduced naive T cell pools are a characteristic feature of aging and may contribute to the impaired immune responses observed in elderly individuals^38^. Depletions in naive T cell populations, and the corresponding deficits in adaptive immune responses, have also been reported in inflammatory states like chronic hepatitis C infection^39^ and chronic granulomatous disease^40^; however, this phenomenon has been less well documented in solid organ transplantation. Additionally, it has been consistently observed that solid organ transplant recipients develop poor adaptive response to new antigens either during immunization or new infections – including SARS-CoV-2 mRNA vaccination^41–43^. It is unlikely that the patient’s immunosuppression prior to primary infection led to naive T cell specific lymphopenia as MMF, a purine biosynthesis inhibitor, would be expected to inhibit both T and B cell proliferation non-specifically, and this patient, having received Belatacept, a CTLA-4 Fc fusion protein and co-stimulatory inhibitor, would also be expected to inhibit T cell activation and differentiation, which would more likely lead to increased, rather than decreased, naive T cell pools. Additionally, cytokine profiling revealed high levels of IL-7 and IL-15, both of which promote naive T cell pool expansion^44,45^. While it is clear that the patient had sufficient cytokines to replenish naive T cell pools, naive T cell populations were not replenished, possibly due to either over-utilization (via repeated antigen engagement), insufficient thymic reserve, or some combination of both. Interestingly, immunophenotyping also revealed high levels of activation, terminal differentiation, and exhaustion in both CD4^+^ and CD8^+^ pools, possibly as a result of chronic antigen exposure from the transplanted organ^46^. We suspect that the lack of naive T cell pools may have contributed to a deficient humoral immune response during initial SARS-CoV-2 infection. Whether similar impaired cellular dynamics may lead to impaired humoral immunity to SARS-CoV-2 in other long-term organ graft recipients, or other populations with aspects of repeated antigen exposure such as chronic infection and cancer, warrants further investigation.

While the patient’s overall anti-SARS-CoV-2 response was not specifically impaired by low naïve CD4 T cell pools, we suspect that the patient’s poor neutralization response may have been and that the insufficient T cell support resulted in either extrafollicular or dysfunctional germinal center B cell responses^34,47–49^. In line with these findings, the patient demonstrated a transient, relatively poor-quality neutralizing antibody response during initial infection consistent with a short-lived extrafollicular response or deficient germinal center dynamics.

SARS-CoV-2 specific IgG antibodies were detected as early as 5 DFSO* during reinfection, suggesting a memory response given the accelerated humoral kinetics relative to first infection. Additionally, the absence of circulating B cell and antiviral IgM during the reinfection indicates that it was likely only SARS-CoV-2 specific plasma cells established after the primary infection that provided neutralizing antibodies during reinfection. To better characterize these neutralizing antibodies, we performed linear epitope profiling of SARS-CoV-2 specific antibodies from the primary infection and reinfection against the spike protein. This revealed binding peaks at identical amino acid locations, suggesting that a humoral memory response was indeed generated during the patient’s primary infection and was also present during reinfection. Importantly, the spike protein amino acid change we identified in the virus strain causing the SARS-CoV-2 reinfection did not correspond to the location of antibody binding generated during the primary infection, suggesting viral evasion of primary humoral responses to be an unlikely explanation for reinfection in this case (**Fig. 5f, Extended Data Fig. S2**). These two binding sites within the spike protein corresponded to amino acid 141 and 1112, which did not correspond to areas of high antigenicity or neutralization with in our larger IMPACT cohort (**Extended Data Fig. S6**), We hypothesize that the patient’s underlying immune deficiencies (low naïve CD4 pools) led to poor neutralizing antibody quality (IC50 titers approximately 1:10 to 1:30), which were insufficient to protect against SARS-CoV-2 reinfection *in vivo*.

By means of a case study of a solid organ transplant recipient with COVID-19, we demonstrate that the mere presence of neutralizing antibodies during primary infection was insufficient to confer protection against reinfection. Further investigation into additional immunological correlates of protection, including the roles of cellular immunity and tissue-resident immune cell populations, are warranted.

### Limitations of Study

As with all case studies, a limitation on the generalizability of our findings to wider patient populations is present. Also, while the lack of immunological responsiveness to vaccination or acute infection in immunosuppressed and solid organ transplant populations is well documented, there may be additional mechanisms contributing to these defects beyond those discussed in this manuscript - particularly with regards to SARS-CoV-2 infection. Our analysis of the immunophenotype of the patient was limited to surveys of circulating immune dynamics; however, numerous studies have also described perturbations in immunity at tissue sites not easily amenable to direct interrogation. We also did not directly analyze antigen specific T-cell responses during either infection, which may reveal additional dysfunction not discussed within this manuscript. Lastly, we also did not fully address every potential avenue of viral immune evasion to immune responses and accordingly suggest that a greater understanding of the virus-intrinsic and host-intrinsic features determining susceptibility to SARS-CoV-2 reinfections is required. Future studies should investigate not only the circulating and systemic adaptive immune responses during SARS-CoV-2 reinfections, but also the possibility that local defects in immune responsiveness among barrier tissue sites may also enable recurrent SARS-CoV-2 infection.

### Quantification and Statistical Analysis

All statistical analysis was performed using R and MATLAB software. Specific statistical methods and results are reported in relevant figures legends and Methods sections.

## Data Availability

Correspondence and requests regarding clinical features of this manuscript should be addressed to M.A. Other correspondence and requests should be addressed to A.I. and B.I.

## Supplemental Information

Supplementary information can be accessed at:

## Acknowledgements

We thank M. Linehan for technical and logistical assistance. We thank C. Wilen for kindly providing the SARS-CoV-2 virus. We also give special recognition of the services of B. Fontes, D. Scoville, and the Yale EH&S Department for their on-going assistance in safely conducting biosafety level 3 research. This work was supported by the Women’s Health Research at Yale Pilot Project Program (A.I.), Fast Grant from Emergent Ventures at the Mercatus Center (A.I. and N.D.G), Mathers Foundation, the Ludwig Family Foundation, the Department of Internal Medicine at the Yale School of Medicine, Yale School of Public Health, the Beatrice Kleinberg Neuwirth Fund, and CDC Contract # 75D30120C09570 (N.D.G.). IMPACT received support from the Yale COVID-19 Research Resource Fund. A.I. is an Investigator of the Howard Hughes Medical Institute. J.K. received support from NIH T32 MSTP training grant. B.I received support from NIAID 2T32AI007517-16. T.A. and M.E.P were supported by CTSA Grant Number TL1 TR001864.

## Author Contributions

A.I., B.I., M.A., and N.D.G conceived the study. J.K., A.F.B., P.T., M.A, B.I., and A.I. drafted the manuscript. P.T., B.I., and M.A constructed the patient’s clinical narrative. T.A., C.B.F.V., J.R.F., A.W.L, M.E.P, C.C.K, and IMPACT Team members performed RNA extractions, PCR, and virus sequencing from patient samples. A.F.B. performed phylogenetic analysis. J.K., C.L., T.Y., J.O., J.S., and IMPACT Team members performed sample collection and PBMC isolation. P.W. and P.L. performed flow cytometry and analyzed data. F.L. performed ELISA assays on patient samples. J.K., M.P.N., and C.L. performed PRNT50 assays. All authors helped to edit the manuscript. A.I. and N.D.G. secured funds and supervised the project.

## Extended Data

**Extended Data Table S1.**
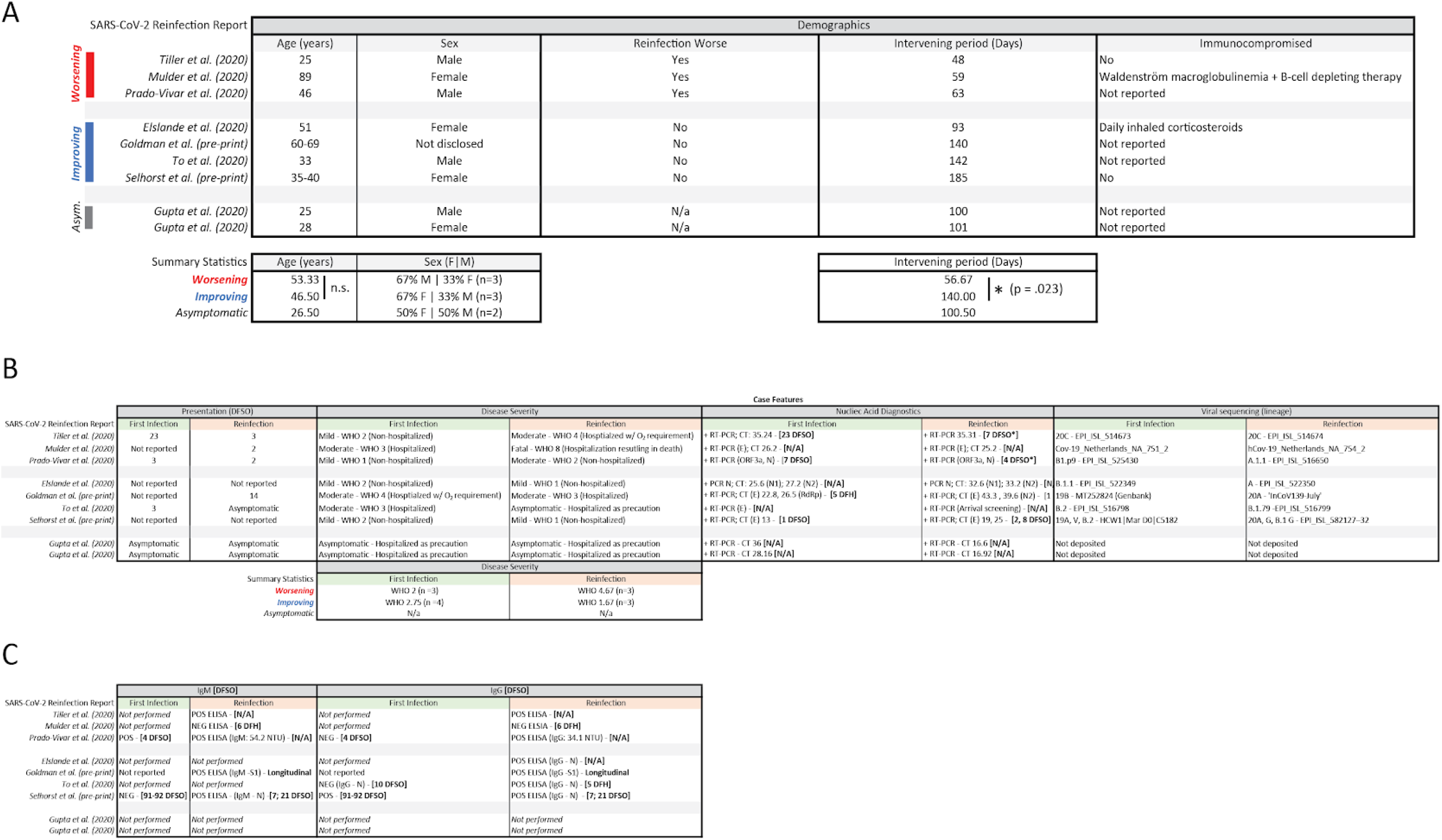
Literature summary of SARS-CoV-2 reinfection reports. **(A)** Demographic and clinical features of SARS-CoV-2 reinfection reports, stratified by clinical progression (worsening, improving, asymptomatic) and ordered by length of intervening period between infections (shortest first). Summary statistics are grouped below in corresponding columns. Group averages between ‘Worsening’ and ‘Improving’ were compared for significance using a two-samples t-test, with equal variances between groups for ‘Age’, and unequal variances for ‘Intervening Period’ as determined by results from F-test statistics. **(B)** Corresponding case features for SARS-CoV-2 reinfection reports stratified by primary infection (green) and reinfection (orange). DFSO represents reported days from symptom onset for primary (green) and reinfection (orange) cases. World Health Organization (WHO) COVID-19 disease severity was assigned retrospectively during the course of this review and were not supplied by the original authors. Where available, nucleic acid results are reported as: Type of test (nucleic acid target): CT values [DFSO or days from hospitalization (DFH)]. Accession numbers for viral genomes are reported as provided in manuscripts. **(C)** Serological results for SARS-CoV-2 reinfection reports stratified by primary infection (green) and reinfection (orange). IgM and IgG data are presented as reported in manuscripts according to the following scheme: Type of test / Result of Test: Quantitative value [DFSO or DFH].

**Extended Data Table S2:**
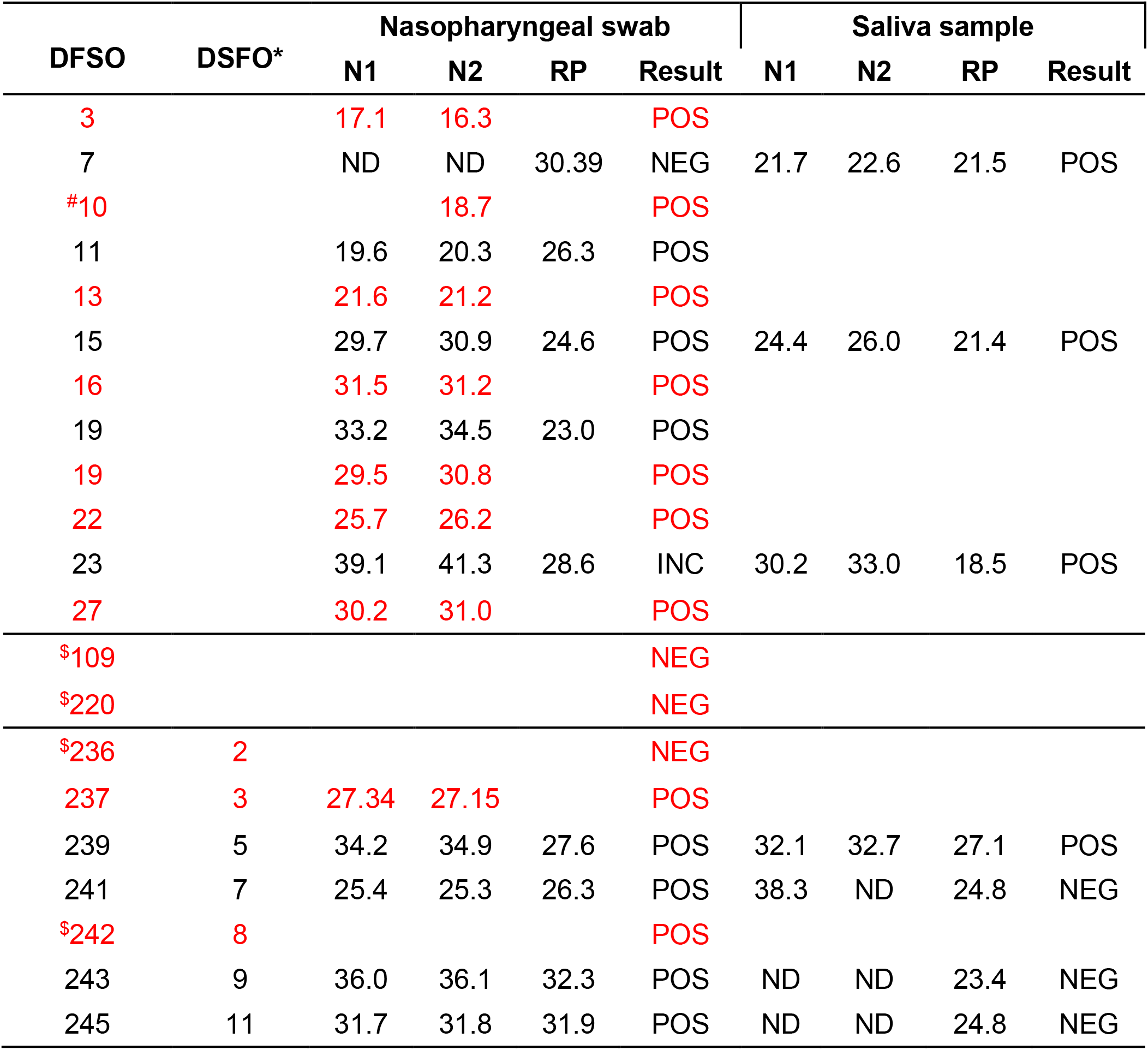
RT-qPCR results for nasopharyngeal swabs and saliva specimens. DFSO corresponds to days from symptom onset during the patient’s primary SARS-CoV-2 infection, whereas DFSO* corresponds to days from symptom onset relative to the SARS-CoV-2 reinfection. Results of nucleic acid testing from each day and for each sample type are reported in consecutive rows ordered by DFSO. N1 and N2 columns correspond to CDC-N1 and CDC-N2 primer sets for the detection of SARS-CoV-2 nucleic acids; RP serves as a positive control for RNA extraction. RT-qPCR cycle threshold (CT) values are reported for various samples, with “ND” indicating SARS-CoV-2 nucleic acids were not detected. As previously described^50^, summary test results are stratified by CDC-N1 CT values according to the following scheme: positive (“POS”) = CT ≤ 38, negative (“NEG”) = CT ≥ 40, or Inconclusive (“INC”) = CT ≥ 38 and CT ≤ 40. Black text indicates samples processed by research lab. Red text indicates samples processed by clinical lab. ND= not detected. # run on Cepheid platform. $ run on Panther platform.

**Extended Data Figure S1.**
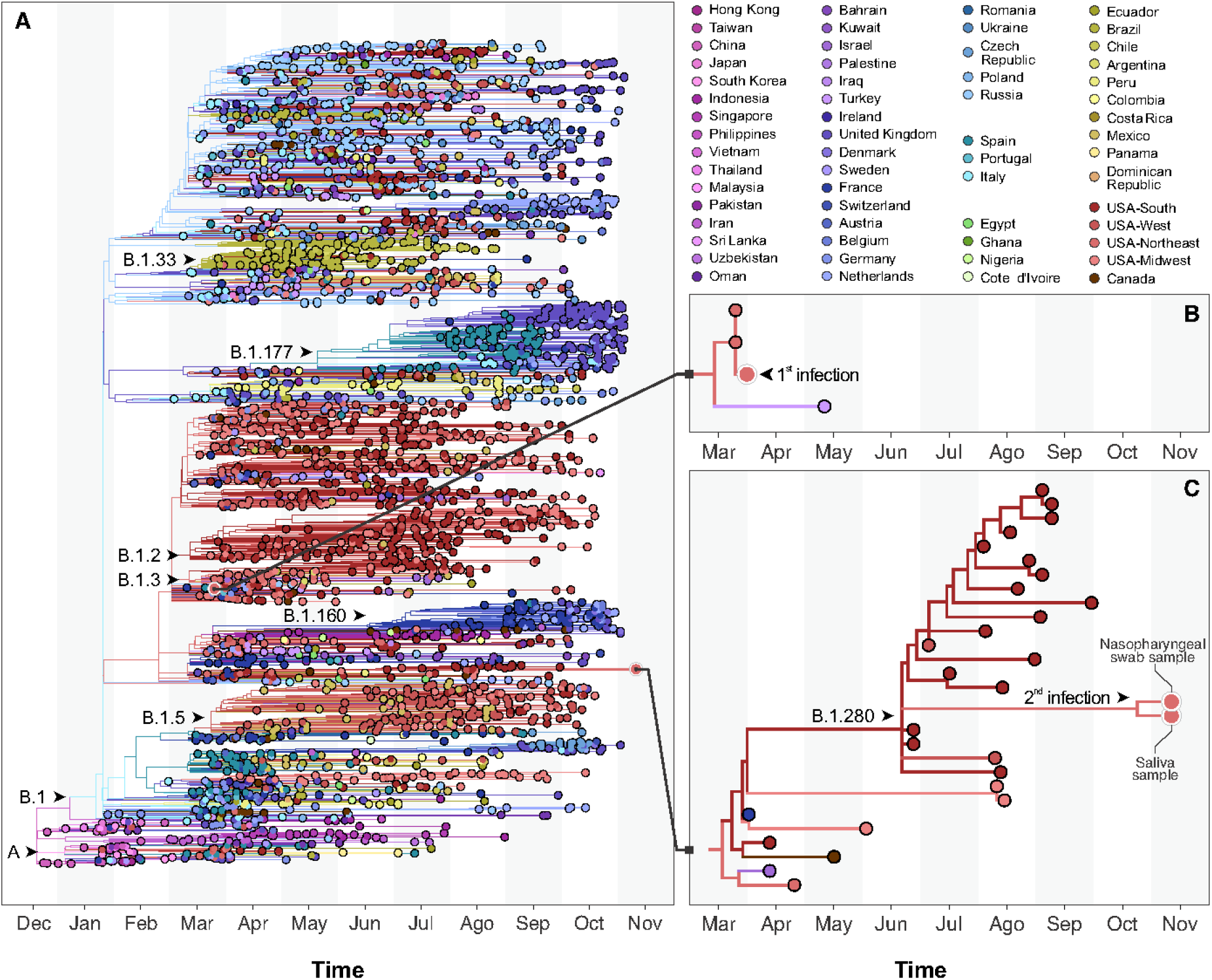
Large phylogeny of SARS-CoV-2 whole genomes. **(A)** Maximum likelihood tree reconstructed with 3,068 global viral genomes from GISAID, sampled to represent the diversity of viruses in circulation since the beginning of the COVID-19 pandemic. This phylogeny revealed the placement of the viruses infecting the patient in distinct lineages/sub-lineages of SARS-CoV-2. **(B)** The virus causing the primary infection was isolated in March 2020 and belongs to lineage B.1. **(C)** The virus causing the reinfection was collected in November 2020 and is found in the sublineages B.1.208. Phylogenetic plots were done using baltic 0.1.0.

**Extended Data Figure S2.**
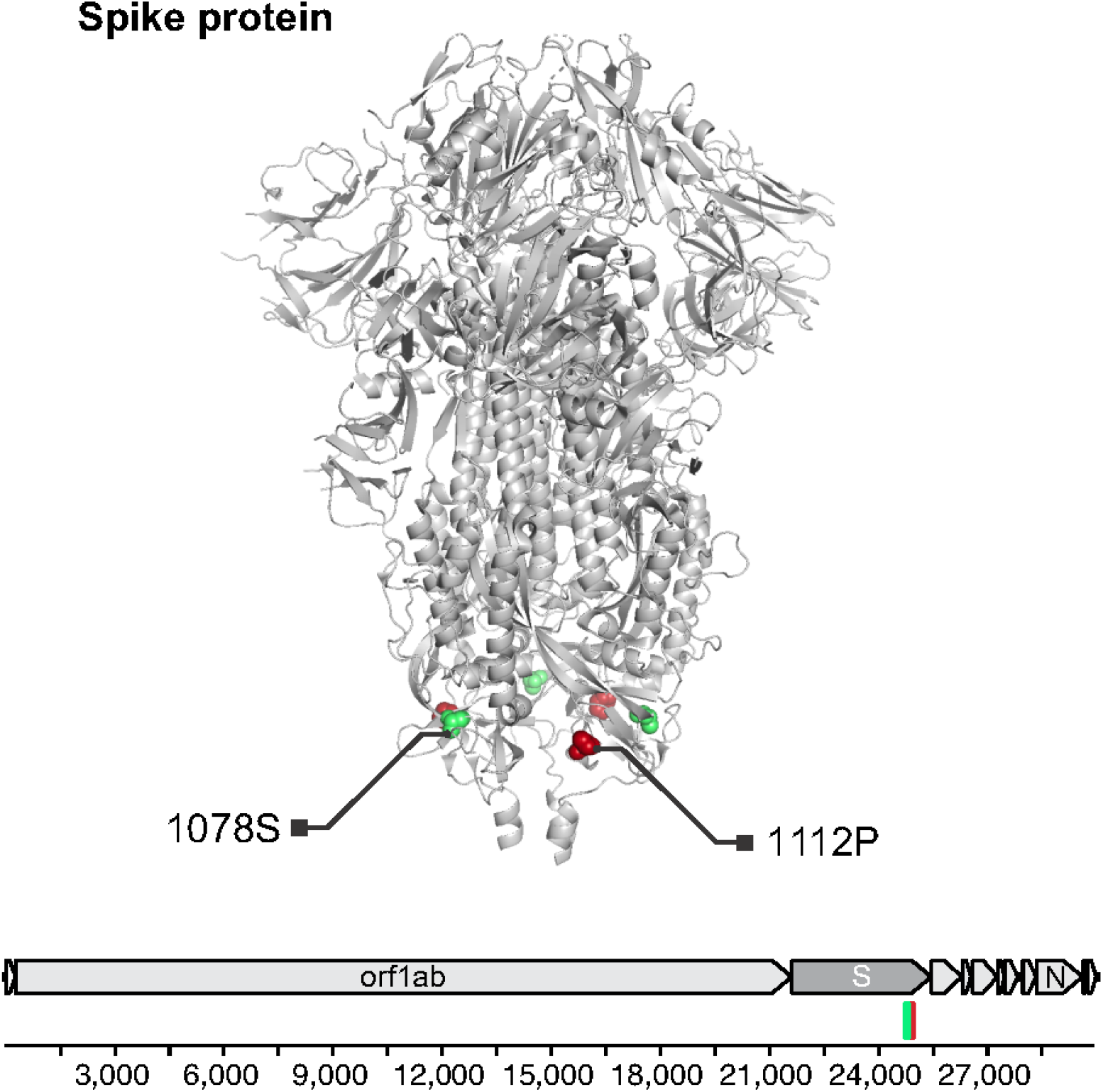
A non-synonymous amino acid change in the genome of the SARS-CoV-2 reinfection variant. An amino acid change from alanine to serine (A1078S) was discovered in the S2 subunit of spike protein in the SARS-CoV-2 variant that caused the reinfection (green marker). Antibody binding site identified by linear epitope mapping that was generated during primary infection (red). Protein structure: PDB 6VXX^51^

**Extended Data Figure S3.**
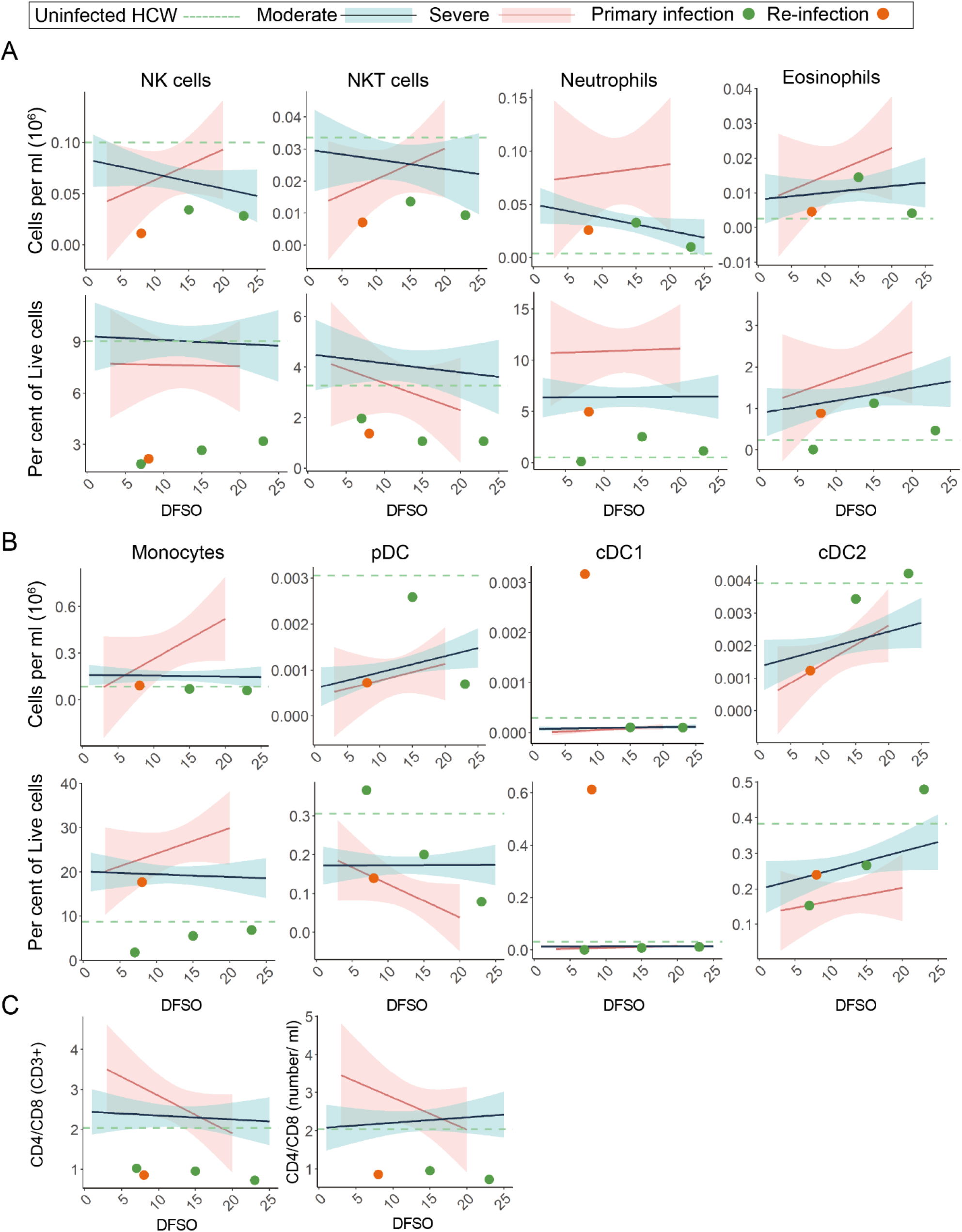
Broad PBMC profiling of SARS-CoV-2 primary and reinfection. For all graphs, blue linear least squares regression lines and corresponding shading represent the average trend and error bars, respectively, for patients with moderate COVID-19. Red linear least squares regression lines and corresponding shading represent the average trend and error bars, respectively, for patients with severe COVID-19. The dashed green line represents the average value of healthy, uninfected healthcare workers (HCW) plotted as a constant value across all days for reference. Individual scatter points represent the values for the patient during the primary SARS-CoV-2 infection (green) and the reinfection (orange). **(A)** Scatter plots of various patient peripheral NK and granulocyte cell populations isolated from whole blood. Top rows are absolute counts, bottom rows are percentage of relative parent populations. **(B)** Scatter plots of various patient peripheral monocyte and dendritic cell (DC) sub-populations isolated from whole blood. Top rows are absolute counts, bottom rows are percentage of relative parent populations. **(C)** CD4^+^/CD8^+^ ratios plotted as percentage of CD3^+^ PMBC cells (left) or as a ratio of absolute counts (right).

**Extended Data Figure S4.**
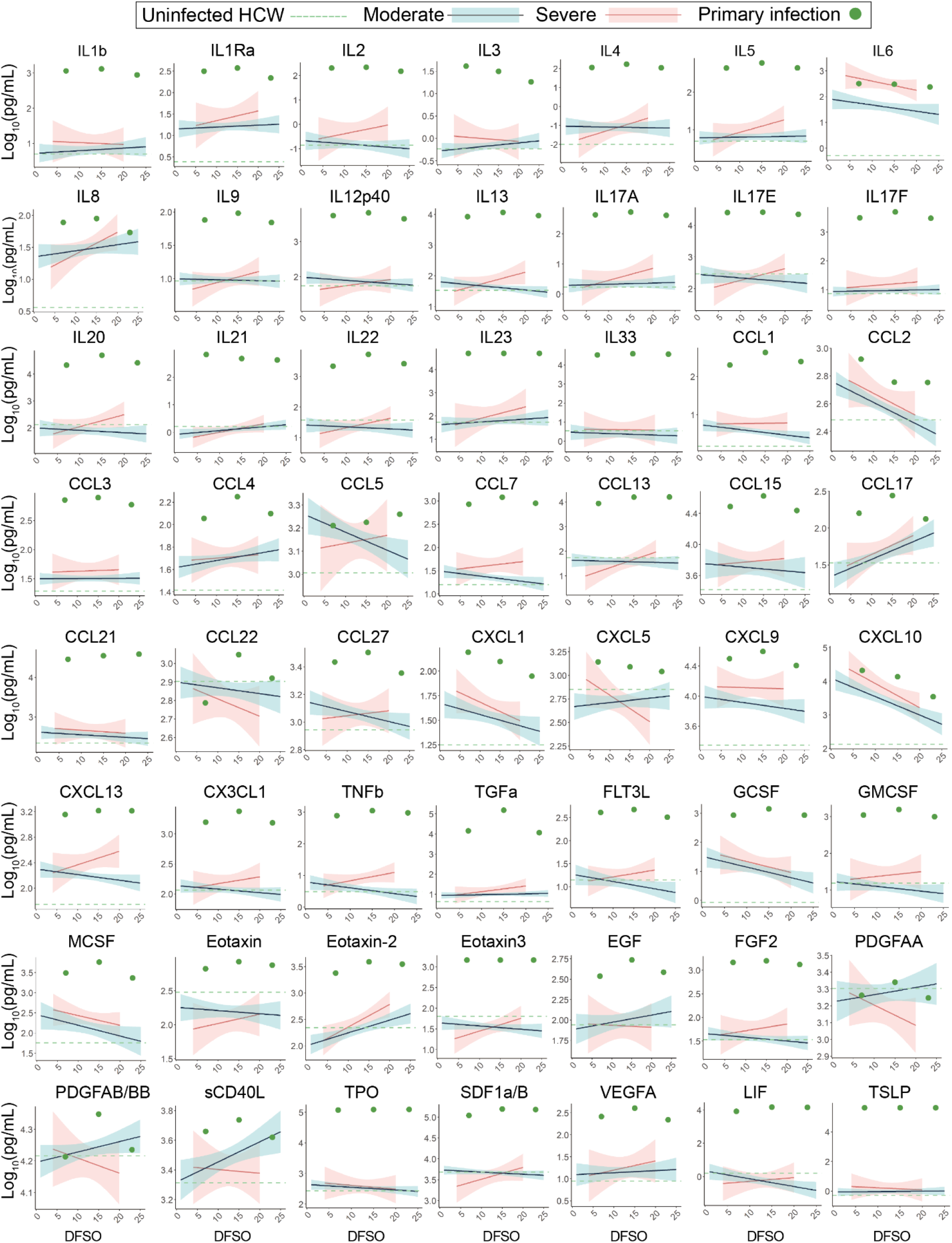
Broad plasma cytokine profiling of SARS-CoV-2 primary infection. For all graphs, blue linear least squares regression lines and corresponding shading represent the average trend and error bars, respectively, for patients with moderate COVID-19. Red linear least squares regression lines and corresponding shading represent the average trend and error bars, respectively, for patients with severe COVID-19. The dashed green line represents the average value of healthy, uninfected healthcare workers (HCW) plotted as a constant value across all days for reference. Individual scatter points represent the values for our patient during the primary SARS-CoV-2 infection (green).

**Extended Data Figure S5.**
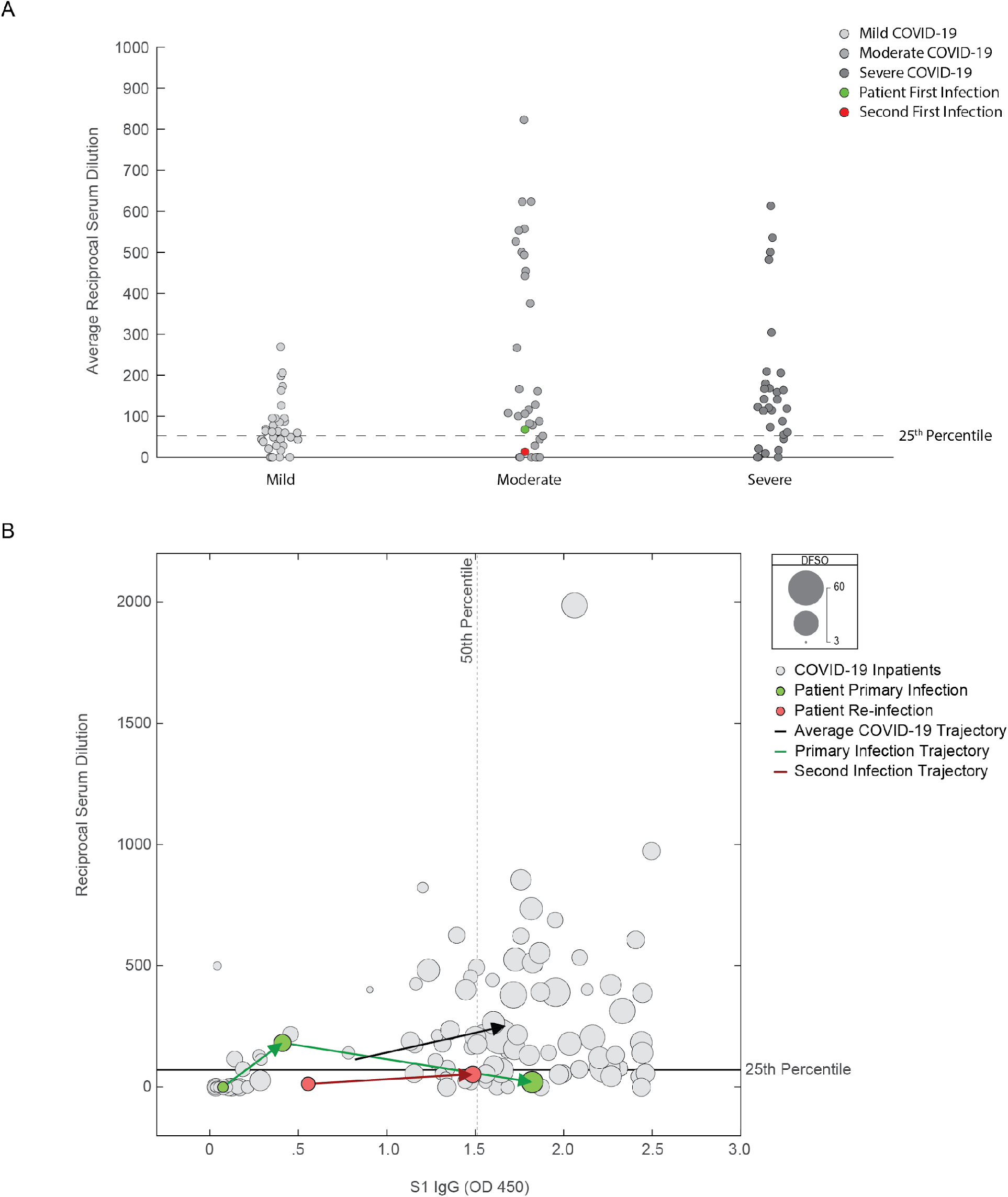
Patient’s suboptimal humoral responses as compared to a cohort of COVID-19 patients. **(A)** Primary infection (green) and reinfection (orange) reciprocal IC_50_ serum dilutions plotted against control COVID-19 patients and stratified by COVID-19 disease severity. Grey points indicate the average reciprocal serum dilution for an individual patient. Dashed line represents the 25^th^ percentile value among patients with COVID-19 who mounted a neutralizing antibody response. **(B)** Reciprocal serum dilution values plotted against corresponding S1 IgG values for each sample collected. Scatter size is scaled against days from symptom onset for each patient. Dashed line indicates the 50^th^ percentile value of IgG response, whereas the solid line represents the 25^th^ percentile reciprocal serum dilution IC_50_ value among patients that mounted a neutralizing antibody response. Green arrows indicate the temporal progression from sequential samples during the primary infection. Orange arrows indicate the temporal progression from sequential samples during the reinfection. Black arrow represents the average trajectory of humoral responses for a control COVID-19 cohort.

**Extended Data Figure S6.**
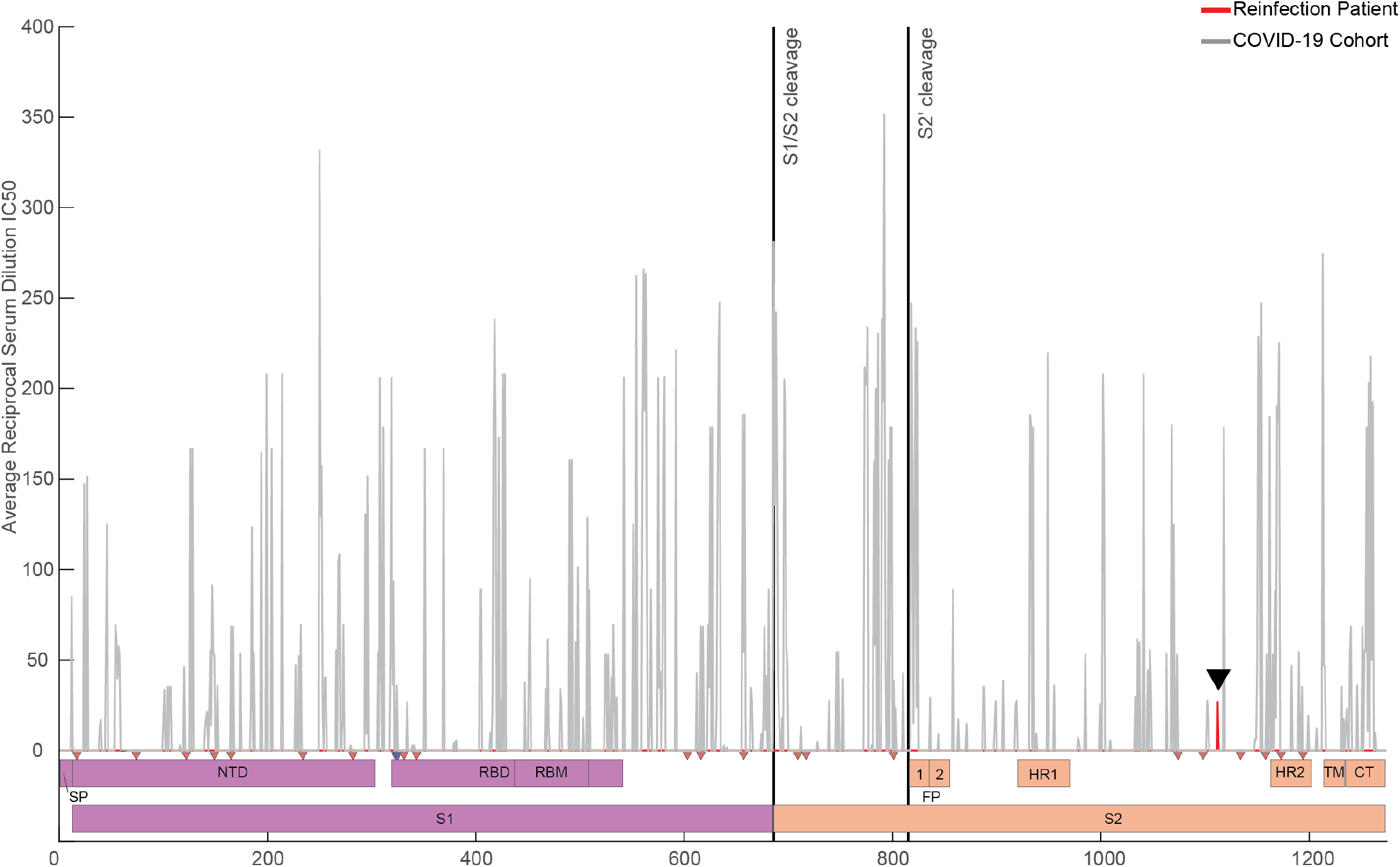
Neutralizing antibody landscape of the SARS-CoV-2 spike protein. Reciprocal serum dilution IC_50_ values are plotted according to identified PIWAS peak locations for hospitalized patients with COVID-19 in the IMPACT cohort (grey). All reciprocal serum dilution values at a given residue were averaged to generate a cohort average value per residue of spike protein. Red peak (black arrow at residue 1112) represents the averaged reciprocal serum dilution IC_50_ for the patient’s primary and reinfections (Peak at 140 not shown). **(Bottom)** Domain map of SARS-CoV-2 spike protein showing S1 (purple) and S2 (orange) regions. N-Glycosylation sites are marked by red triangles, O-linked glycosylation sites are marked by blue triangles. Cleavage sites are marked by solid black lines (“S1/S1 Cleavage” and “S2’ cleavage”).

## Supplementary Information

**Supplementary Table S1. List of genomes used in the analyses**. We gratefully acknowledge the following Authors from the Originating laboratories responsible for obtaining the specimens, as well as the submitting laboratories where the genome data were generated and shared via GISAID, on which this research is based.

**Supplementary Table S2. Yale IMPACT Research Team**

Abeer Obaid^10^, Adam J. Moore^2^, Alice Lu-Culligan^1^, Allison Nelson^10^, Angela Nunez^10^, Anjelica Martin^1^, Anne E. Watkins^2^, Bertie Geng^10^, Christina A. Harden^2^, Codruta Todeasa^10^, Cole Jensen^2^, Daniel Kim^1^, David McDonald^10^, Denise Shepard^10^, Edward Courchaine^11^, Elizabeth B. White^2^, Eric Song^1^, Erin Silva^10^, Eriko Kudo^1^, Giuseppe DeIuliis^10^, Harold Rahming^10^, Hong-Jai Park^10^, Irene Matos^10^, Isabel Ott^2^, Jessica Nouws^10^, Jordan Valdez^10^, Joseph Lim^12^, Kadi-Ann Rose^10^, Kelly Anastasio^13^, Kristina Brower^2^, Laura Glick^10^, Lokesh Sharma^10^, Lorenzo Sewanan^10^, Lynda Knaggs^10^, Maksym Minasyan^10^, Maria Batsu^10^, Maxine Kuang^2^, Maura Nakahata^10^, Melissa Campbell^6^, Melissa Linehan^1^, Michael H. Askenase^14^, Michael Simonov^10^, Mikhail Smolgovsky^10^, Nicole Sonnert^1^, Nida Naushad^10^, Pavithra Vijayakumar^10^, Rick Martinello^3^, Rupak Datta^3^, Ryan Handoko^10^, Santos Bermejo^10^, Sarah Prophet^15^, Sean Bickerton^11^, Sofia Velazquez^14^,Tyler Rice^1^, William Khoury-Hanold^1^, Xiaohua Peng^10^, Yexin Yang^1^, Yiyun Cao^1^, Yvette Strong^10^

^10^Yale School of Medicine, New Haven, CT, USA.

^11^Department of Biochemistry and of Molecular Biology, Yale University School of Medicine, New Haven, CT, USA.

^12^Yale Viral Hepatitis Program; Yale University School of Medicine, New Haven, CT, USA.

^13^Yale Center for Clinical Investigation, Yale University School of Medicine, New Haven, CT, USA.

^14^Department of Neurology, Yale University School of Medicine, New Haven, CT, USA.

^15^Department of Molecular, Cellular and Developmental Biology, Yale University School of Medicine, New Haven, CT, USA.

## METHODS Legend

**KEY RESOURCES TABLE**

**RESOURCE AVAILABILITY**

Lead Contacts

Materials Availability

Data Availability

**EXPERIMENTAL MODEL AND SUBJECT DETAILS**

Ethics statement

Patient Case Report and IMPACT Patient Cohort

Cell lines

SARS-CoV-2 Virus

**METHOD DETAILS**

Isolation of patient plasma and PBMCs

SARS-CoV-2 RT-qPCR detection and sequencing

Phylogenetic and molecular evolution analysis

Flow cytometry

SARS-CoV-2 specific-antibody measurements

Enzyme-Linked Immunosorbent Assay (ELISA)

Cytokine and chemokine measurements

Viral neutralization assay

STAR METHODS Text

### RESOURCE AVAILABILITY

#### Lead Contacts

Correspondence and requests regarding clinical features of this manuscript should be addressed to M.A. Other correspondence and requests should be addressed to A.I.

#### Materials availability

Requests materials should be addressed to B.I. and A.I.

#### Data availability

Requests for data or should be addressed to B.I. and A.I.

### EXPERIMENTAL MODEL AND SUBJECT DETAILS

#### Ethics statement

This study was approved by Yale Human Research Protection Program Institutional Review Boards (FWA00002571, protocol ID 2000027690). Informed consent was obtained from all enrolled patients and healthcare workers. All work requiring biosafety level three conditions was performed by certified personnel under the supervision of the Yale Department of Environmental Health and Safety.

#### Patient Case Report and IMPACT Patient Cohort

Our case report of a SARS-CoV-2 reinfection was enrolled as part of the broader Yale IMPACT cohort described at length previously and summarized again below for convenience^52,53^. The individual clinical narrative for our SARS-CoV-2 reinfection case was written by two infectious disease fellows and an infectious disease physician specializing in management of solid organ transplant recipients through direct interview of the patient and review of their EMR. All explicit dates in our clinical narrative were converted to relative days from symptom onset as reported by our patient. With regards to the larger IMPACT cohort, and reproduced here for convenience, one hundred and seventy-nine patients admitted to YNHH with COVID-19 between 18 March 2020 and 27 May 2020 were included in this study. No statistical methods were used to predetermine sample size. Patients were scored for COVID-19 disease severity through review of the electronic health records (EMR) at time of each sample collection. Scores were assigned by a clinical infectious disease physician according to a custom-developed disease severity scale. Moderate disease status (clinical scores 1–3) was defined as: (1) SARS-CoV-2 infection requiring hospitalization without supplementary oxygen; (2) infection requiring non-invasive supplementary oxygen (<3 l/min to maintain SpO2 >92%); and (3) infection requiring non-invasive supplementary oxygen (>3 l/min to maintain SpO2 >92%), or alternatively >2 l/min to maintain SpO2 >92% with a high-sensitivity C-reactive protein (hsCRP) >70 and administration of tocilizumab. Severe disease status (clinical score 4 or 5) was defined as: (4) infection meeting all criteria for clinical score 3 and also requiring admission to the ICU and >6 l/min supplementary oxygen to maintain SpO2 >92%, or (5) infection requiring invasive mechanical ventilation or extracorporeal membrane oxygenation (ECMO) in addition to glucocorticoid or vasopressor administration. Clinical score 6 was assigned for deceased patients. For all patients, days from symptom onset were estimated as follows: (1) highest priority was given to explicit onset dates provided by patients; (2) next highest priority was given to the earliest reported symptom by a patient; and (3) in the absence of direct information regarding symptom onset, we estimated a date through manual assessment of the EHR by an independent clinician. Symptom onset and etiology were recorded through standardized interviews with patients or patient surrogates upon enrollment in our study, or alternatively through manual EHR review if no interview was possible. Clinical data were collected using EPIC EHR, and de-identified and aggregated using REDCap 9.3.6 software.

## METHOD DETAILS

### Isolation of patient plasma and PBMCs

As reported previously and reproduced here for convenience^52,53^, patient whole blood was collected in sodium heparin-coated vacutainers and kept gently agitating at room temperature until sample pick-up by IMPACT team members. All blood was processed on the day of collection. Plasma samples were collected after centrifugation of whole blood at 400*g* for 10 minutes at room temperature (RT) without brake. The undiluted serum was then transferred to 15-ml polypropylene conical tubes, and aliquoted and stored at −80 °C for subsequent analysis. PBMCs were isolated using Histopaque (Sigma-Aldrich, #10771-500ML) density gradient centrifugation in a biosafety level 2+ facility. After isolation of undiluted serum, blood was diluted 1:1 in room temperature PBS, layered over Histopaque in a SepMate tube (StemCell Technologies; #85460) and centrifuged for 10 minutes at 1,200*g*. The PBMC layer was isolated according to the manufacturer’s instructions. Cells were washed twice with PBS before counting. Pelleted cells were briefly treated with an ACK lysis buffer for 2 minutes and then counted. Percentage viability was estimated using standard Trypan blue staining and an automated cell counter (Thermo-Fisher, #AMQAX1000).

### SARS-CoV-2 RT-qPCR detection and sequencing

Total nucleic acid was extracted from nasopharyngeal swabs and saliva specimens with the MagMAX Viral/Pathogen Nucleic Acid Isolation kit^54^. We used the modified CDC RT-qPCR assay to detect SARS-CoV-2 in the extracted nucleic acid^50,55^. A selection of specimens from both time periods that tested positive were sequenced using a highly multiplexed amplicon sequencing approach as described in the ARTIC Network nCoV-2019 Oxford Nanopore sequencing protocol <u>(nCoV-2019 sequencing protocol v3 (LoCost) V.3)^56^</u>. Sequencing data were then processed according to the ARTIC bioinformatic pipeline. Briefly, raw .fast5 data was basecalled with Guppy V.4.4.0 and aligned to the reference SARS-CoV-2 genome (Accession MN908947) using minimap2. Primer sequences are masked prior to consensus sequence generation. A threshold of 20X depth of coverage is required to call variants and generate a consensus sequence. Positions in the genome with less than 20X depth of coverage are represented with “N”.

### Phylogenetic and molecular evolution analysis

To reveal the evolutionary relationship between the virus infecting the patient in March 2020 with those sampled during the reinfection in November 2020, phylogenetic analyses were performed with additional sequences submitted to GISAID (gisaid.org), sampled from all over the world from January to mid-November 2020. Metadata on viral collection time, location, and lineage assignment were also obtained from GISAID. A time series of daily COVID-19 cases per country, obtained from the Center for Systems Science and Engineering (CSSE) data repository at Johns Hopkins University^57^, was used to weight the genome subsampling according to the daily case counts. This allowed a balanced sampling of 3,068 genomes out of nearly 265,000 available on GISAID up to November 19th, 2020 (**Extended Data Table S1**), which ensured more accurate discrete phylogeographic reconstruction to unravel the likely origins of viruses from the 1st and 2nd infections. The genome sequences were aligned using MAFFT^58^, and the inference of the global phylogeny (n = 3,068 genomes) was performed using IQTree v.1.6.12, applying a GTR substitution model^59^, and TreeTime v.0.8.0^60^ within an augur pipeline^61^. From this large preliminary phylogeny (**Extended Fig. S1**), the expected clustering of the virus genomes sequenced from the patient was verified, and the sequence dataset was further subsampled, to include only 561 genomes belonging to closely related SARS-CoV-2 sublineages (**Extended Fig. S1**). The same approach implemented in the first round of analysis was used to generate the final phylogeny, but ultrafast bootstrap (1000 replicates)^62^ was performed to assess branch support. Phylogenetic data visualization was done using auspice (interactive view)^61^ and baltic 0.1.0 (static view). The search for polymorphisms in comparison with the reference genome (MN908947.3) was performed using Python scripts, and visualization of genome annotations were done using DNA Features Viewer 3.0.1^63^. The structure in Extended Data Fig. S2 (PDB 6VXX) was downloaded from PDB^51^, and formatted using PyMol. (The PyMOL Molecular Graphics System, Version 2.0 Schrödinger, LLC.)

### Flow cytometry

As detailed in previous manuscripts and reproduced here for convenience^52^, antibody clones and vendors used for flow cytometry analysis were as follows: BB515 anti-hHLA-DR (G46-6) (1:400) (BD Biosciences), BV785 anti-hCD16 (3G8) (1:100) (BioLegend), PE-Cy7 anti-hCD14 (HCD14) (1:300) (BioLegend), BV605 anti-hCD3 (UCHT1) (1:300) (BioLegend), BV711 anti-hCD19 (SJ25C1) (1:300) (BD Biosciences), AlexaFluor647 anti-hCD1c (L161) (1:150) (BioLegend), biotin anti-hCD141 (M80) (1:150) (BioLegend), PE-Dazzle594 anti-hCD56 (HCD56) (1:300) (BioLegend), PE anti-hCD304 (12C2) (1:300) (BioLegend), APCFire750 anti-hCD11b (ICRF44) (1:100) (BioLegend), PerCP/Cy5.5 anti-hCD66b (G10F5) (1:200) (BD Biosciences), BV785 anti-hCD4 (SK3) (1:200) (BioLegend), APCFire750 or PE-Cy7 or BV711 anti-hCD8 (SK1) (1:200) (BioLegend), BV421 anti-hCCR7 (G043H7) (1:50) (BioLegend), AlexaFluor 700 anti-hCD45RA (HI100) (1:200) (BD Biosciences), PE anti-hPD1 (EH12.2H7) (1:200) (BioLegend), APC anti-hTIM3 (F38-2E2) (1:50) (BioLegend), BV711 anti-hCD38 (HIT2) (1:200) (BioLegend), BB700 anti-hCXCR5 (RF8B2) (1:50) (BD Biosciences), PE-Cy7 anti-hCD127 (HIL-7R-M21) (1:50) (BioLegend), PE-CF594 anti-hCD25 (BC96) (1:200) (BD Biosciences), BV711 anti-hCD127 (HIL-7R-M21) (1:50) (BD Biosciences), BV421 anti-hIL17a (N49-653) (1:100) (BD Biosciences), AlexaFluor 700 anti-hTNFa (MAb11) (1:100) (BioLegend), PE or APC/Fire750 anti-hIFNy (4S.B3) (1:60) (BioLegend), FITC anti-hGranzymeB (GB11) (1:200) (BioLegend), AlexaFluor 647 anti-hIL-4 (8D4-8) (1:100) (BioLegend), BB700 anti-hCD183/CXCR3 (1C6/CXCR3) (1:100) (BD Biosciences), PE-Cy7 anti-hIL-6 (MQ2-13A5) (1:50) (BioLegend), PE anti-hIL-2 (5344.111) (1:50) (BD Biosciences), BV785 anti-hCD19 (SJ25C1) (1:300) (BioLegend), BV421 anti-hCD138 (MI15) (1:300) (BioLegend), AlexaFluor700 anti-hCD20 (2H7) (1:200) (BioLegend), AlexaFluor 647 anti-hCD27 (M-T271) (1:350) (BioLegend), PE/Dazzle594 anti-hIgD (IA6-2) (1:400) (BioLegend), PE-Cy7 anti-hCD86 (IT2.2) (1:100) (BioLegend), APC/Fire750 anti-hIgM (MHM-88) (1:250) (BioLegend), BV605 anti-hCD24 (ML5) (1:200) (BioLegend), BV421 anti-hCD10 (HI10a) (1:200) (BioLegend), BV421 anti-CDh15 (SSEA-1) (1:200) (BioLegend), AlexaFluor 700 Streptavidin (1:300) (ThermoFisher), BV605 Streptavidin (1:300) (BioLegend). In brief, freshly isolated PBMCs were plated at 1–2 × 10^6^ cells per well in a 96-well U-bottom plate. Cells were resuspended in Live/Dead Fixable Aqua (ThermoFisher) for 20 min at 4 °C. Following a wash, cells were blocked with Human TruStan FcX (BioLegend) for 10 min at RT. Cocktails of desired staining antibodies were added directly to this mixture for 30 min at RT. For reinfection stains, cells were first washed and supernatant aspirated; then to each cell pellet a cocktail of secondary markers was added for 30 min at 4 °C. Prior to analysis, cells were washed and resuspended in 100 μl 4% PFA for 30 min at 4 °C. Following this incubation, cells were washed and prepared for analysis on an Attune NXT (ThermoFisher). Data were analysed using FlowJo software version 10.6 software (Tree Star). The specific sets of markers used to identify each subset of cells are summarized in Extended Data Fig. 9 of Lucas et al. (2020)^33^.

### SARS-CoV-2 specific-antibody measurements

ELISAs were performed as previously described and reproduced here for convenience^52,64^. Briefly, Triton X-100 and RNase A were added to serum samples at final concentrations of 0.5% and 0.5mg/ml respectively and incubated at room temperature (RT) for 30 minutes before use to reduce risk from any potential virus in serum. 96-well MaxiSorp plates (Thermo Scientific #442404) were coated with 50 μl/well of recombinant SARS Cov-2 S1 protein (ACROBiosystems #S1N-C52H3-100ug) at a concentration of 2 μg/ml in PBS and were incubated overnight at 4 °C. The coating buffer was removed, and plates were incubated for 1h at RT with 200 μl of blocking solution (PBS with 0.1% Tween-20, 3% milk powder). Serum was diluted 1:50 in dilution solution (PBS with 0.1% Tween-20, 1% milk powder) and 100 μl of diluted serum was added for two hours at RT. Plates were washed three times with PBS-T (PBS with 0.1% Tween-20) and 50 μl of HRP anti-Human IgG Antibody (GenScript #A00166, 1:5,000) or anti-Human IgM-Peroxidase Antibody (Sigma-Aldrich #A6907, 1:5,000) diluted in dilution solution added to each well. After 1 h of incubation at RT, plates were washed three times with PBS-T. Plates were developed with 100 μl of TMB Substrate Reagent Set (BD Biosciences #555214) and the reaction was stopped after 15 min by the addition of 2 N sulfuric acid. Plates were then read at a wavelength of 450 nm and 570nm.

### Cytokine and chemokine measurements

Patient serum was isolated as already described in this manuscript and previously reported^52^. Briefly, aliquots were stored at −80 °C. Sera were shipped to Eve Technologies (Calgary, Alberta, Canada) on dry ice, and levels of cytokines and chemokines were measured using the Human Cytokine Array/Chemokine Array 71-403 Plex Panel (HD71). All samples were measured upon the first thaw.

### Cell lines and virus

As previously reported^65^, Vero E6 kidney epithelial cells were cultured in Dulbecco’s Modified Eagle Medium (DMEM) supplemented with 1% sodium pyruvate (NEAA) and 5% fetal bovine serum (FBS) at 37°C and 5% CO2. The cell line was obtained from the ATCC and has been tested negative for contamination with mycoplasma. SARS-CoV-2, strain USA-WA1/2020, was obtained from BEI Resources (#NR-52281) and was amplified in Vero E6 cells. Cells were infected at a MOI 0.01 for four three days to generate a working stock and after incubation the supernatant was clarified by centrifugation (450g × 5min) and filtered through a 0.45-micron filter. The pelleted virus was then resuspended in PBS then aliquoted for storage at − 80°C. Viral titers were measured by standard plaque assay using Vero E6 cells. All experiments were performed in a biosafety level 3 with the Yale Environmental Health and Safety office approval.

### Neutralization assay

As reported in previous manuscripts and reiterated for convenience^65^, patients and healthy donor sera were isolated from whole blood and heat treated for 30 minutes at 56 °C. Sixfold serially diluted plasma, from 1:10 to 1:2430 were incubated with SARS-CoV-2 for 1 h at 37 °C. The mixture was subsequently incubated with Vero E6 cells in a 6-well plate for 1 hour for infection. Cells were overlayed with MEM supplemented NaHCO3, 2% FBS, and 0.6% Avicel mixture. Plaques were resolved at 40h post infection by fixing in 10% formaldehyde for 1h followed by staining in 0.5% crystal violet. All experiments were performed in parallel with healthy, uninfected control sera to establish degree of protection as reflected in difference in plaque counts.

